# Validating HIV viral suppression threshold adjustments for comparable estimates using data from nationally representative household surveys in sub-Saharan Africa

**DOI:** 10.1101/2025.04.09.25325517

**Authors:** Olanrewaju Edun, Lucy Okell, Timothy M. Wolock, Eline L. Korenromp, Leigh F. Johnson, Jeffrey W. Imai-Eaton

**Author notes:** Corresponding author: Olanrewaju Edun. TMW is currently employed by Schonfeld Strategic Advisors LLC.

## Abstract

**Introduction:** To enable comparable global assessments of viral load suppression (VLS) among people living with HIV on antiretroviral therapy (ART), UNAIDS applies a model to adjust VLS estimates reported at different thresholds to a common VL ≤1000 copies/mL definition. We assessed performance of the current reverse Weibull model and alternatives using survey data from sub-Saharan Africa.

**Methods:** Using data from 21 Population-based HIV Impact Assessment surveys (PHIAs) in 16 sub-Saharan African countries (2015–2022), we assessed six models (Weibull, reverse Weibull, Pareto, Fréchet, gamma, and lognormal) in adjusting VLS reported at VL <50, <200, <400 copies/mL to ≤1000. We compared predictions using parameters from Johnson *et al.* and recalibrated using PHIAs, assessing whether new shape parameters improved adjustments and varied by sex and age.

**Results:** In adjustments from all thresholds, the Weibull model had the lowest prediction errors (Root-mean-squared error for <200 to ≤1000: Weibull: 1.9%; reverse Weibull: 3.1%; Pareto: 2.5%). Prediction errors for reverse Weibull and Pareto models were higher in subgroups with low VLS, compared to Weibull. Across 21 surveys, in adjustments from <200 to ≤1000, reverse Weibull overestimated VLS by 2.3%, compared to 1.5% by Weibull and Pareto. The Fréchet, gamma, and lognormal models performed similarly to Weibull. Shape parameter estimates for the Weibull and reverse Weibull were slightly higher after recalibration and varied by sex and age.

**Conclusion:** The Weibull, Fréchet, gamma, and lognormal models, provided more reliable VLS adjustments across thresholds than the previously recommended reverse Weibull model, avoiding inflated VLS estimates which could obscure gaps in HIV treatment programmes and underestimate HIV transmission risks.

## Introduction

Monitoring HIV viral load (VL) among people living with HIV (PLHIV) is important for assessing antiretroviral therapy (ART) effectiveness and HIV transmission risk (1). The UNAIDS ‘95-95-95’ targets aim for 95% of PLHIV on ART to achieve viral load suppression (VLS) by 2025 (‘third 95’ target), defined as VL ≤1000 copies/mL (2,3). However, some national programmes use different thresholds (<400, <200, or <50 copies/mL) in national monitoring (4–7), leading to spurious differences in VLS across settings or even within a single setting if not accounted for (8–11).

To standardize global monitoring of progress towards the ‘third 95’ target, UNAIDS apply a mathematical formula to adjust VLS estimates from country-reported thresholds to the common VL ≤1000 threshold (8). The adjustment is based on analysis by Johnson *et al.* that fitted Weibull, Pareto, and reverse Weibull models to viral load distribution data from PLHIV on ART in the International epidemiology Databases to Evaluate AIDS (IeDEA) collaboration and the ART Cohort Collaboration (ART-CC) (8). The IeDEA and ART-CC data included PLHIV from seven global regions, including sub-Saharan Africa, and Europe from 2010 to 2019 (8). The calibrated models were validated by assessing their reliability in adjusting VLS estimates using World Health Organization (WHO) drug resistance report data (from Guatemala, Honduras, Nicaragua, Vietnam and Zambia) and Brazil’s national ART programme (8). While Pareto and reverse Weibull models best fit the calibration data (assessed using the log likelihood statistics), the Weibull model performed best in validation with the WHO data (8), though all three model predictions fell within uncertainty limits of the validation data. The Pareto model performed best on validation with the Brazilian programme data, but its uncertainty ranges were the narrowest (8). Given the Weibull’s relatively poorer goodness-of-fit to calibration data and the Pareto’s narrow uncertainty ranges, the study recommended the reverse Weibull model for adjusting estimates of VLS, which UNAIDS incorporated into the Spectrum model in 2021 (8). For example, an observed 80% VLS at a threshold of <200 copies/mL from routine VL monitoring would be adjusted to 88.3% Click or tap here to enter text.at ≤1000 copies/mL using the reverse Weibull model (8).

Due to the limited availability of individual-level VL data, the models were calibrated to aggregate VL data, which may have limited their ability to assess individual-level variability and extreme values, influencing shape parameter estimates. Data were limited to evaluate differences in the VL distributions among individuals by age and sex (8). Also, due to limited data, the performance of the models in adjusting VLS estimates were validated mostly using data from South and central America, with limited validation against data from sub-Saharan Africa, where rates of HIV drug resistance may be lower (12).

Using nationally representative Population-based HIV Impact Assessment surveys (PHIAs) (13) from sub-Saharan Africa, we aimed to (1) evaluate the models estimated by Johnson *et al.* in African populations; (2) compare alternative statistical models for describing observed VL distributions; and (3) assess sex and age differences in VL distribution parameters among PLHIV on ART in sub-Saharan Africa.

## Methods

### Adjustment approaches

Johnson *et al.* explored three models—Weibull, reverse Weibull, and Pareto—to represent the cumulative distribution of VL on the log_10_ scale (log_10_-VL) among PLHIV on ART (8). Table 1 summarizes the cumulative distribution functions (CDFs) for each model. The reverse Weibull model imposed an upper limit of 6 log_10_-VL (1,000,000 copies/mL) on VLs among PLHIV on ART, modelling the difference between an individual’s log_10_-VL and this upper limit using the Weibull CDF (8). Johnson *et al.* estimated shape parameters from empirical cumulative distribution functions (reported in Table 1). Then for a fixed shape parameter, a VLS observation at threshold t_l_ (e.g. <50, <200, <400) was converted to threshold t_2_ (≤1000 copies/mL) by calculating the scale parameter (formula in Table 1). All three models were calibrated using data from the IeDEA and ART-CC cohorts to obtain shape estimates and validated using data form the WHO drug resistance report and the Brazilian national ART programme (8).

**Table 1.**
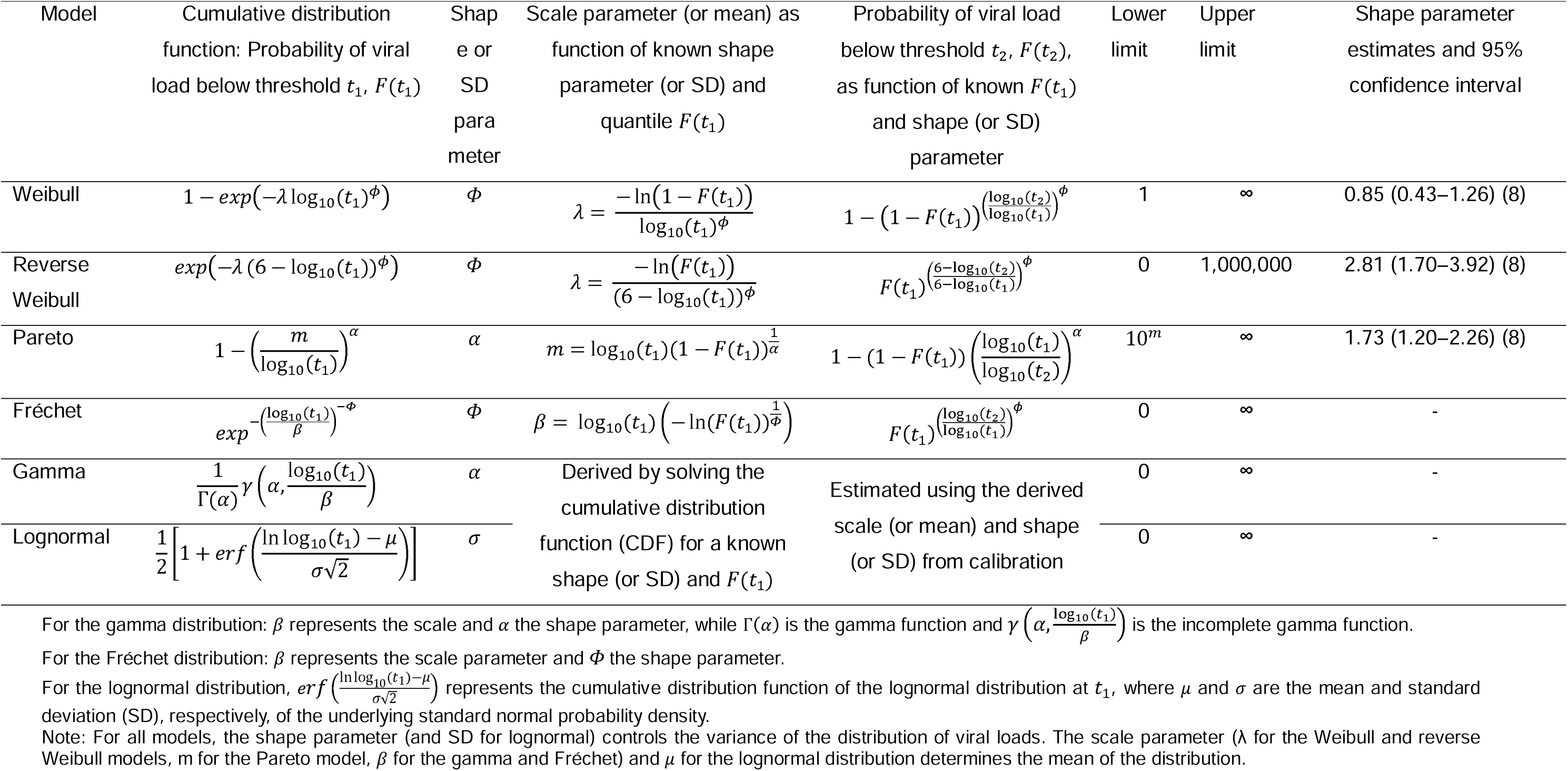
Models of viral load distributions among PLHIV on ART (adapted from Johnson *et al.*).

### Data source

We analysed participant-level VL data from 21 PHIAs conducted in 16 sub-Saharan African countries between 2015–2022. These cross-sectional household surveys collected standardised, nationally representative HIV-related data (13). Surveys included two each from Eswatini, Malawi, Lesotho, Zambia, and Zimbabwe, and one survey from Cameroon, Côte d’Ivoire, Nigeria, Ethiopia, Kenya, Rwanda, Uganda, Tanzania, Namibia, Mozambique, and Botswana.

Details of the PHIA design, sampling and procedures have been described previously (14,15). Consenting participants completed structured questionnaires including demographic information and self-reported ART use. Whole blood samples were collected via venous draw for household-based and laboratory testing. HIV status was determined using point-of-care serological testing, with positive samples confirmed by the Geenius HIV-1/2 Confirmatory Assay (Bio-Rad, Hercules, CA), followed by HIV RNA VL testing and antiretroviral (ARV) screening to assess ART use. (14,15).

Ethical approvals were obtained from institutional review boards in each country, the U.S. Centers for Disease Control and Prevention (CDC), and Columbia University or the University of Maryland at Baltimore (Nigeria, Botswana, and Zambia 2020 surveys). Written informed consent was obtained from adult participants, with assent and parental consent required for minors (typically 15–17 years). This secondary analysis received ethical approval from the Imperial College Research Governance and Integrity Team (ICREC #20IC6451).

Our analyses included participants aged 15 years and older, HIV seropositive, and on ART based on either presence of detectable ARVs in blood or self-reported ART usage.

### Statistical analyses

First, to evaluate the models derived by Johnson *et al.*, we analysed the observed survey-weighted proportions of PLHIV on ART with VL <50, <200 and <400 copies/mL in each survey. These values were then input into the cumulative distributions for the Pareto, Weibull, and reverse Weibull modelsClick or tap here to enter text. (Table 1) to predict the proportion with VL ≤1000 copies/mL. We compared these to the observed survey proportion with VL ≤1000, quantified prediction errors using the root-mean-squared error (RMSE), and reported the average difference between the predicted and observed proportions as a measure of bias. Lower RMSE indicated smaller prediction errors.

Second, we refit the Weibull and reverse Weibull models to individual-level log_10_-VL data from the 21 PHIAs, assuming a common shape parameter for all surveys while allowing country- and survey-specific random effects in the scale parameter. VL measurements reported at the lower limit of quantification (LLOQ) varied across and within surveys and were treated in the likelihood as censored observations at the reported LLOQ (e.g., <20 or <839 copies/mL). Measurements reported as “Target not detected”, without a specified LLOQ, were censored at the lowest quantifiable VL within the survey (ranging from 20 to 40 copies/mL) (Table S1) (16–18). To facilitate fitting the reverse Weibull model, which imposes an upper limit of 6 log_10_-VL, we truncated VL values ≥6 log_10_-VL at 5.99 log_10_-VL for 52 eligible participants with recorded VL above this threshold. We assessed model fits by comparing empirical VL histograms (log_10_ transformed) with probability density estimates of the Weibull, reverse Weibull and Pareto distributions, using both Johnson *et al.*’s parameters and those estimated from PHIA data (8). We assessed whether the recalibrated models improved VLS predictions at ≤1000 using the reported proportions <50, <200 and <400 copies/mL by comparing RMSE and average bias.

Since the Pareto distribution’s scale parameter *m* defines its lower limit, it was unsuitable for likelihood-based fitting to individual-level survey observations. Instead, we tested a range of plausible shape parameter values for the Pareto distribution, visually assessed their fits to PHIA data, and evaluated their predictive performance for VLS ≤1000 copies/mL (Figures S2-S4).

Third, we explored three additional distributions used for modelling right-skewed data, Fréchet, gamma, and lognormal (19–22). Their CDFs are in Table 1. These models were fitted to individual-level log_10_-VL data from the 21 PHIAs as described previously to obtain a common shape parameter. The fixed shape parameter and a VLS observation at threshold *t*_l_ were then used to convert to a threshold *t*_2_ (≤1000 copies/mL) by calculating the scale parameter, as described previously (Table 1). We then compared their performance against the Johnson *et al.* models using the RMSE and average bias.

Lastly, to assess if best fitting shape parameters differed by age and sex, we fit the models to PHIA data using distributional regression, in which the shape or standard deviation parameters vary with respect to covariates (23). We applied distributional regression, specifying the shape parameter as a log-linear function of sex and age categories (15–24, 25–34, 35–44, 45–54 and 55+ years), including country- and survey-specific random effects for the scale parameter. We then compared VLS predictions using these sex- and age-specific parameters to those from sex- and age-invariant parameters, using RMSE and average bias (8).

Adjustments from <200 to ≤1000 copies/mL were used to illustrate results, as most countries reporting VLS estimates to UNAIDS at alternative thresholds use <200 copies/mL (Table S2). Analyses were conducted in R version 4.3.1, with model calibration using the *brms* package (version 2.20.4).

## Results

Across 21 surveys, VL measurements were available for 36,368 PLHIV on ART (age 15+ years; maximum age eligibility varied). The observed percentage virally suppressed (VL ≤1000 copies/mL) ranged from 73.7% in Côte d’Ivoire (2017–2018) to 97.9% in Botswana (2021) (Table S1).

### Evaluation of the Johnson et al. models using PHIA data

Using shape parameters estimated from Johnson *et al.* (8) to predict VL ≤1000 copies/mL from observed proportions <50, <200 and <400 copies/ml, all three original models overestimated the proportion ≤1000 in most PHIAs (Figures 1 and S1). The average bias for predicted VL ≤1000 adjusting from <50, <200 and <400 for the reverse Weibull model was 1.3%, 2.3% and 1.6%, respectively. Average bias was 0.4%, 1.5% and 0.8% for the Weibull model and 2.0%, 1.5% and 0.8% for the Pareto (Table S3). However, predicted VLS proportions were within survey uncertainty ranges, except for adjustments from <50 for Botswana (2021), where all three models significantly underestimated VLS.

**Figure 1.**
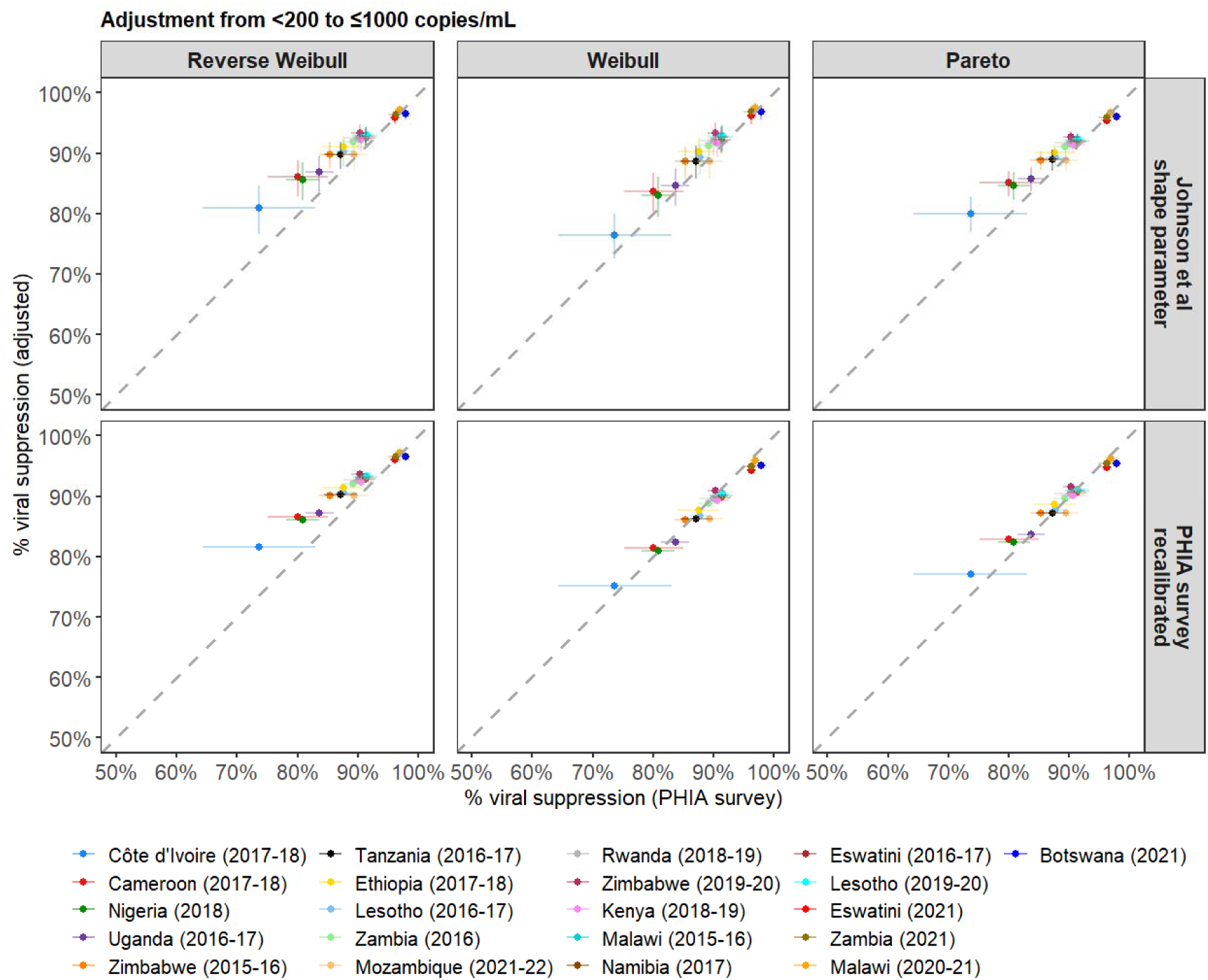
Scatter plots show the relationship between the observed percentage VLS estimates from the individual patient data in the PHIAs and the adjusted estimates (from <200 to ≤1000 copies/mL) using the reverse Weibull, Weibull and Pareto models and parameters from Johnson et al and calibration to PHIA survey data. Shape parameter for the Pareto model on calibration to PHIA survey data used in Figure is 1.20 (see Figure S3 for comparisons using other values). The surveys are arranged in ascending order based on the observed proportion of VLS at the 1000 copies/mL threshold.

The Weibull model had the smallest RMSE across adjustments: from <50 (3.0%), <200 (1.9%), and <400 (1.1%), compared to reverse Weibull (3.2%, 3.1%, 2.1%) and Pareto (3.8%, 2.5%, 1.4%). The reverse Weibull and Pareto models had larger prediction errors for surveys with lower VLS (Figures 1 and S1).

After recalibration to PHIAs, the shape parameter for the Weibull (0.91; 95% CI: 0.90–0.92) and reverse Weibull (2.98; 2.93–3.02) were slightly higher but similar to Johnson *et al.* (Weibull: 0.85; 0.43–1.26 and reverse Weibull: 2.81; 1.70–3.92) (8). The recalibrated Weibull model improved RMSE from 1.9% to 1.4% for predictions using observed proportion <200, and from 1.1% to 0.8% for <400, but worsened from 3.0% to 4.3% for <50 (Table S3). The recalibrated reverse Weibull model slightly worsened predictions across all three thresholds, with larger prediction errors for surveys with lower levels of VLS. On average, it overestimated VL<1000 by 1.9%, 2.6% and 1.8% in adjustment from <50, <200 and <400, respectively, while the recalibrated Weibull underestimated VLS by 3.4%, 0.8% and 0.4%, respectively.

For Pareto, the best-fitting shape parameter varied by VLS level. Higher values (≥2.00) fit better for VLS >90%, while lower values (<2.00) for VLS <90% (Figure S2). Prediction errors for VL ≤1000 copies/mL were higher with shape ≥2.00 than with Johnson *et al.’*s shape estimate (1.73), especially in lower-VLS surveys (Figure S3). A lower shape (1.20) reduced errors. RMSE for adjustment from <200 to ≤1000 was 1.4% (shape = 1.20), 2.5% (1.73), 3.1% (2.00), and 4.2% (2.50) (Figure S3; Appendix S1; Figure S4).

Comparisons of fitted model densities and empirical VL distributions showed all models underestimated the upper tail (VL >1000 copies/mL), especially in surveys with VLS <90% (Figure 2A–C). The reverse Weibull and Pareto underestimated this more than the Weibull model, particularly with PHIA-calibrated shape estimates (Figures 2A–D).

**Figure 2.**
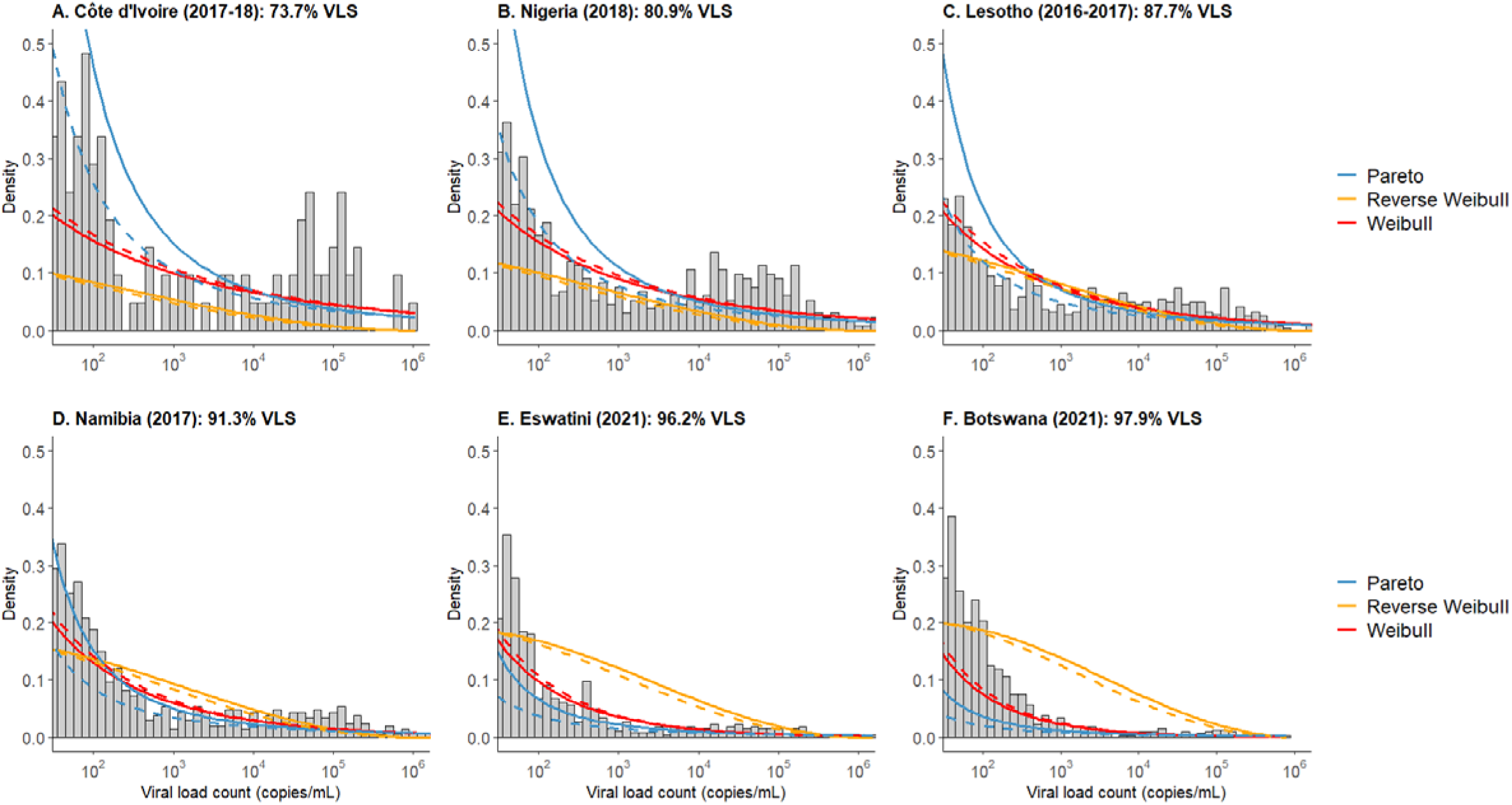
Histograms show the distribution of observed viral loads among PLHIV on ART in the A. Côte d’Ivoire (2017-18); B. Côte 263 d’Ivoire (2017-18); B. Nigeria (2018); C. Lesotho (2016-2017); D. Namibia (2018); E. Eswatini (2021) and F. Botswana (2021) PHIA surveys. Lines show the probability density estimates for the Pareto, reverse Weibull and Weibull models using shape parameters from Johnson et al. (solid line) and calibration to PHIA surveys (dashed lines). The dashed line for the Pareto model is for shape = 1.20 (see Figure S2 for other values). Note: the scale parameters were set so the cumulative probability of a viral load ≤1000 copies/mL is the same as VLS estimated from the survey data. The x-axis was truncated at 50 copies/mL for visualization purposes. Gaps in the histogram for Côte d’Ivoire reflect sparse data points for certain viral load values on the x-axis due to the small number of PLHIV on ART (n = 207).

In surveys with VLS>95%, all three models underestimated VL density <1000 copies/mL (Figures 2E–2F). Pareto and Weibull underestimated low VL values more than the reverse Weibull model, in these surveys. Increasing the shape parameter for Pareto and reverse Weibull reduced the underestimation for VL <1000 copies/mL in high-VLS surveys (Figure S2).

### Fréchet, gamma and lognormal models

Table 2 reports the best fitting shape and standard deviation parameters for these models. None improved predictions of VL ≤1000 copies/mL over the Weibull model using Johnson *et al*.’s or PHIA recalibrated parameters, which performed best (Figures 3, and S5). RMSE for Fréchet, gamma and lognormal models were lower than for reverse Weibull and Pareto models (using Johnson *et al.*’s or recalibrated parameters) but similar to Weibull (Table 2). Plots of the VL density fits for the Fréchet, gamma and lognormal models (Figure S6) closely match those of the Weibull model.

**Figure 3.**
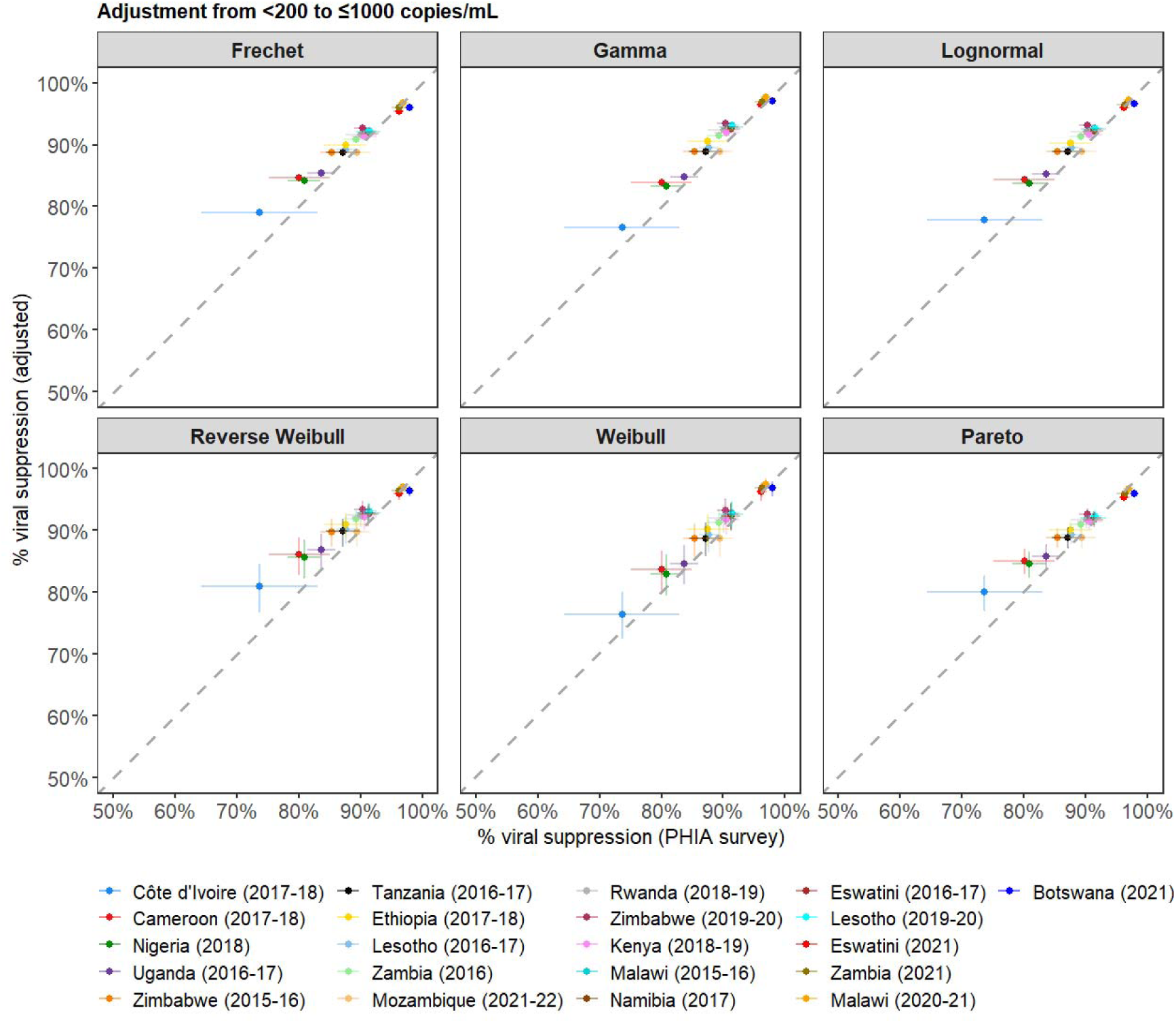
Scatter plots show the relationship between the observed percentage VLS estimates from the individual patient data in the PHIA surveys and the adjusted estimates (from <200 to ≤1000 copies/mL) using the Fréchet, gamma and lognormal model (parameters from calibration to the PHIA surveys), and the reverse Weibull, Weibull and Pareto models (using parameters from Johnson *et al*.). Surveys in legend are sequenced in increasing order of observed VLS.

**Table 2.** Shape parameter estimates and 95% confidence intervals for the reverse Weibull, Weibull, Pareto, Fréchet, gamma, and lognormal models estimated from calibration to the PHIA survey data.

| Model | Johnson *et al.* | Pooled | Men | Women | 15-24 years | 25-34 years | 35-44 years | 45-54 years | 55+ years |
| --- | --- | --- | --- | --- | --- | --- | --- | --- | --- |
| Reverse Weibull | 2.81 (1.70, 3.92) | 2.98 (2.93, 3.02) | 2.86 (2.78, 2.94) | 3.04 (2.94, 3.13) | 2.46 (2.35, 2.58) | 2.52 (2.37, 2.66) | 2.84 (2.68, 3.00) | 3.81 (3.64, 3.99) | 4.32 (4.10, 4.55) |
| Weibull | 0.85 (0.43, 1.26) | 0.91 (0.90, 0.92) | 1.01 (0.98, 1.03) | 0.88 (0.85, 0.90) | 0.55 (0.53, 0.56) | 0.98 (0.97, 0.99) | 0.98 (0.97, 0.99) | 0.99 (0.98, 1.02) | 1.00 (0.98, 1.04) |
| Pareto | 1.73 (1.20, 2.26) | - | - | - | - | - | - | - | - |
| Fréchet | - | 1.86 (1.83, 1.90) | 1.94 (1.88, 1.99) | 1.84 (1.77, 1.90) | 1.52 (1.43, 1.60) | 1.69 (1.58, 1.80) | 1.88 (1.78, 1.98) | 2.13 (2.02, 2.25) | 2.27 (2.13, 2.40) |
| Gamma | - | 0.81 (0.79, 0.84) | 1.02 (0.97, 1.07) | 0.73 (0.69, 0.79) | 0.82 (0.74, 0.90) | 0.73 (0.65, 0.83) | 0.81 (0.73, 0.90) | 0.97 (0.87, 1.08) | 1.01 (0.89, 1.14) |
| Lognormal | - | 0.89 (0.88, 0.90) | 0.83 (0.80, 0.85) | 0.92 (0.90, 0.95) | 0.99 (0.94, 1.03) | 0.96 (0.90, 1.02) | 0.89 (0.83, 0.94) | 0.80 (0.75, 0.86) | 0.78 (0.72, 0.83) |

### Model validation by sex and age

Using Johnson *et al.*’s parameters, prediction errors were slightly higher for men than women for Weibull, reverse Weibull and Pareto models (Figure S7). The Weibull model had the lowest errors in both sexes, except for <50 to ≤1000 adjustment in men, where reverse Weibull performed best (Table S3). On average, in adjustment from <200 to ≤1000 using Johnson *et al.*’s parameters, the reverse Weibull model overestimated VLS by 2.6% (men) and 2.2% (women), Weibull by 1.2% and 1.5%, and Pareto by 1.8% and 1.5%, respectively.

By age, using Johnson *et al.*’s parameters, prediction errors were higher in younger age groups than older ones (Figure S8). The Weibull model consistently had the lowest errors among 15-24 and 25-34 year-olds, while all three models performed similarly for those 35+ (Table S3). For adjustment from <200 to ≤1000, RMSE in 15-24-year-olds was 5.9% (reverse Weibull), 3.0% (Weibull) and 5.3% (Pareto); among those 55+, RMSE were 2.0%, 1.8%, and 1.8%, respectively. On average, VLS was overestimated by 4.6% (reverse Weibull), 1.5% (Weibull), and by 3.7% (Pareto) in the youngest group, but by <1% in those 55+.

Following recalibration to PHIAs, reverse Weibull shape estimates were similar by sex (men: 2.86; 95%CI: 2.78–2.94; women 3.04; 2.94–3.13), but increased with age (Table 2). Weibull shape estimates were higher in men (1.01; 0.98–1.03) than women (0.88; 0.85–0.90), and lower among 15-24-year-olds but stable across older age groups (Table 2).

Using these sex- and age-specific parameters in the reverse Weibull model yielded similar prediction errors to Johnson *et al.*’s parameters for men, women, and individuals 35+ (Table S3). Prediction errors for the reverse Weibull model were slightly lower for those under 35 when using the age-specific shape estimated from the PHIAs. Adjustments using sex- and age-specific parameters the Weibull model produced only marginal changes (Tables 2, S2). For the Pareto model, using a shape of 1.20 reduced errors across most groups, except for those 45-54 and 55+, where errors were similar or higher compared to Johnson *et al.*’s shape estimate.

On calibration to PHIAs, shape parameters for the Fréchet model were similar by sex (men: 1.94; 1.88–1.99; women: 1.84; 1.77–1.90) but varied by age, with higher shape with increasing age (Table 2). Gamma model estimates were higher in men (1.02; 0.97–1.07) than women (0.73; 0.69–0.79), similar among those under 45, and higher in those 45 and older. For the lognormal model, standard deviation was lower in men (0.83; 0.80–0.85) than women (0.92; 0.90–0.95), and decreased with increasing age.

However, applying the Fréchet, gamma, or lognormal model to predict VL ≤1000 copies/mL by sex and age did not improve overall predictions compared to the Weibull (age/sex-indifferent) model (Tables 2, S3-S5).

## Discussion

We found that statistical models proposed by Johnson *et al.* to adjust VLS reported at different thresholds to the global standard of ≤1000 copies/mL overestimated VLS for most national survey from sub-Saharan Africa, although point estimates remained within survey-based uncertainty ranges. Prediction errors were larger when adjusting from lower thresholds (<50) compared to higher thresholds (<200 or <400), and for surveys and subgroups with lower VLS (e.g., men, younger individuals). Of six models explored, the Weibull model had lower prediction errors across all threshold and subgroups than the reverse Weibull and Pareto models, and performed similarly to the Fréchet, gamma and lognormal models. This contrasts the conclusion of Johnson *et al.* to use the reverse Weibull model (8).

The overestimation of VLS by all six models and higher prediction errors in adjustments from lower thresholds is similar to validation results from Johnson *et al.* (8). Likewise, Johnson *et al.* found that shape parameters for the reverse Weibull and Pareto models were correlated with VLS. Specifically, they found higher shape parameter estimates for these models were estimated in regions with higher VLS levels and in adults with higher levels of VLS compared to children (8). Their Weibull estimates were similar across groups, aligning with our findings; however, while Johnson *et al.* examined difference between children and adults, our analysis focused on variation among adult age groups (8). In contrast to our results where prediction errors were lowest for the Weibull model across all adjustments, in Johnson *et al.*, the Weibull model only performed best on validation using the WHO drug resistance data at all thresholds, with similar performance across the three models on validation with the Brazilian national programme data at all thresholds (8). This is related to the high VLS levels (>85%) reported across all timepoints in the Brazilian data (8), whereas in the WHO drug resistance data the Weibull model had the lowest prediction errors for data from Honduras and Nicaragua with VLS of <75% using the ≤1000/mL threshold (8).

Johnson *et al.* noted that viral load distributions among PLHIV on ART can vary substantially even when the fraction virally suppressed is the same (8). We found that the shape of the viral load distribution among PLHIV on ART varied systematically with the fraction virally suppressed, with consistent discrepancies from our model distributions. In settings with high VLS (>95%), the distribution was right-skewed with a long tail, which was suitably modelled using the distributions we explored. In settings with lower VLS (<95%), the distribution had a slight hump of high VL values at the tail, which all explored underestimated. Statistical distributions whose shape or standard deviation parameters were correlated with VLS— reverse Weibull, Pareto, Fréchet and lognormal—required higher shape (or lower standard deviation) parameters to fit high VLS data but the reverse for low VLS settings. This explains why the reverse Weibull model, when calibrated to PHIAs with an average VLS of 89%, produced slightly higher, age-/sex-indifferent shape estimates compared to Johnson *et al*.’s calibration to adult data, where the average VLS was lower (85%). Furthermore, in low VLS settings, the reverse Weibull and Pareto models exhibited poor fit and larger prediction errors when higher shape parameters were used.

Our results suggest that the reverse Weibull model—recommended by the UNAIDS Reference Group on Estimates, Modelling and Projections Click or tap here to enter text.for standardizing viral load measurements (8)—overestimates VLS more than the other five models in this analysis, particularly in low VLS settings. Adjustments using the reverse Weibull model in age/sex subgroups or countries with low VLS may lead to spuriously high levels of VLS, masking existing gaps in subgroups with suboptimal adherence or ART programmes challenges (24). Overestimation of VLS can lead to underestimation of HIV transmission and new infections in mathematical models like the UNAIDS-supported Spectrum model where HIV transmission risk is informed by VLS levels (25).

While the Weibull model outperformed the reverse Weibull and Pareto and was comparable to the Fréchet, gamma or lognormal models at adjusting VLS, it has limitations. First, it also fit poorly in low VLS settings, slightly overestimating VLS, especially for adjustments from the lowest (<50) threshold. Prediction errors for the Weibull model, like other models, were higher when adjusting from <50 to ≤1000 than from <200 or <400 to ≤1000. Conversely, due to its long upper tail, the Weibull model may predict too high viral loads at the upper end of the distribution, limiting its use in modelling HIV transmission from PLHIV on ART (8). While the empirical distribution of VLs among PLHIV on ART in low VLS settings appeared bimodal, we did not explore bimodal or mixture distributions since their parameters would likely correlate with VLS levels and not give simple mathematical adjustments. Although the gamma and lognormal models performed similarly to Weibull, they also lack simple closed-form mathematical adjustments, limiting practical use.

This study has several limitations. First, using data exclusively from sub-Saharan Africa may restrict the generalizability of our shape estimates, particularly for the reverse Weibull, Pareto, Fréchet and lognormal models, to other regions. Second, our calibration relied on survey data, which may limit applicability of our results for adjusting estimates derived from routine programmatic data. Delays in processing routine viral load specimens, as reported in some studies, can lead to inconsistent performance of viral load assays at lower levels of viremia, potentially influencing VL distributions observed in routine programme data (26).

In conclusion, the Weibull model was more reliable for adjusting VLS estimates across thresholds than the recommended reverse Weibull model, which produced worse over-estimation of VLS. Despite its limitations, the Weibull model’s consistent shape parameter throughout most of the range of typical VL distributions, consistency in performance across subgroups, and its mathematically simple adjustment make it the best currently available option for the UNAIDS-supported Spectrum model used for global epidemic estimation and evaluation of progress towards population-level viral load suppression. However, all adjustment methods have the potential to introduce error, and efforts should be made to support countries in reporting using the recommended thresholds, to avoid reliance on adjustment methods.

## Data Availability

PHIA survey data can be requested from the PHIA Project Team at ICAP at Columbia University, except for the Nigeria (2018) survey, which is available upon request from the Nigeria National Bureau of Statistics, and the Botswana (2021) survey, which can be accessed at Statistics Botswana. All other data produced in the present work are contained in the manuscript.

https://phia-data.icap.columbia.edu/datasets

https://microdata.nigerianstat.gov.ng/index.php/catalog/65/related-materials

https://microdata.statsbots.org.bw

## Supplementary information

**Table S1.**
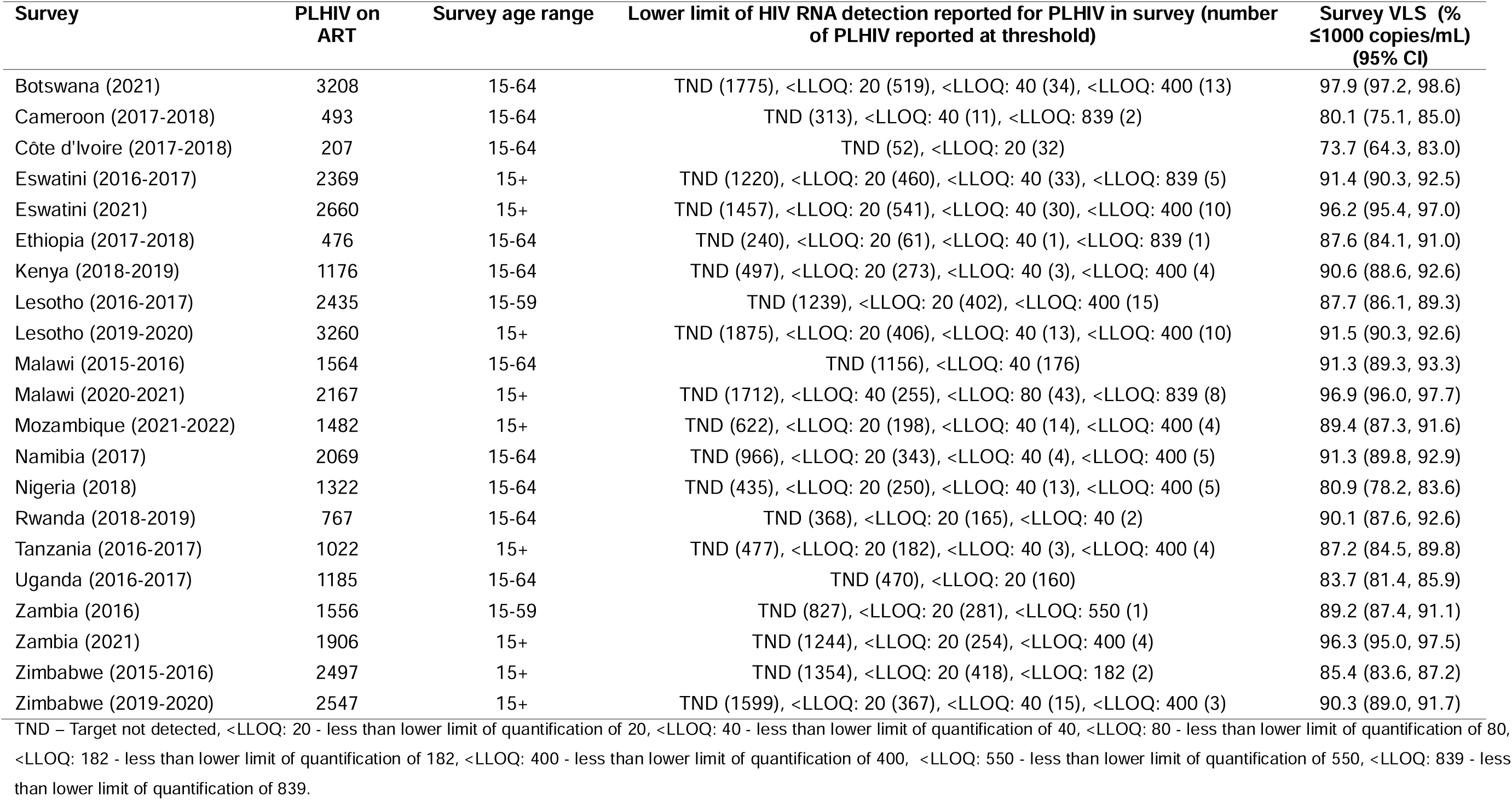
Total number, age range, lower limit of HIV RNA detection and survey weighted VLS estimates and 95% confidence interval for people living with HIV on antiretroviral therapy included from each PHIA survey.

| Survey | PLHIV on ART | Survey age range | Lower limit of HIV RNA detection reported for PLHIV in survey (number of PLHIV reported at threshold) | Survey VLS (% $\leq 1000$ copies/mL) (95% CI) |
| --- | --- | --- | --- | --- |
| Botswana (2021) | 3208 | 15-64 | TND (1775), <LLOQ: 20 (519), <LLOQ: 40 (34), <LLOQ: 400 (13) | 97.9 (97.2, 98.6) |
| Cameroon (2017-2018) | 493 | 15-64 | TND (313), <LLOQ: 40 (11), <LLOQ: 839 (2) | 80.1 (75.1, 85.0) |
| Côte d'Ivoire (2017-2018) | 207 | 15-64 | TND (52), <LLOQ: 20 (32) | 73.7 (64.3, 83.0) |
| Eswatini (2016-2017) | 2369 | 15+ | TND (1220), <LLOQ: 20 (460), <LLOQ: 40 (33), <LLOQ: 839 (5) | 91.4 (90.3, 92.5) |
| Eswatini (2021) | 2660 | 15+ | TND (1457), <LLOQ: 20 (541), <LLOQ: 40 (30), <LLOQ: 400 (10) | 96.2 (95.4, 97.0) |
| Ethiopia (2017-2018) | 476 | 15-64 | TND (240), <LLOQ: 20 (61), <LLOQ: 40 (1), <LLOQ: 839 (1) | 87.6 (84.1, 91.0) |
| Kenya (2018-2019) | 1176 | 15-64 | TND (497), <LLOQ: 20 (273), <LLOQ: 40 (3), <LLOQ: 400 (4) | 90.6 (88.6, 92.6) |
| Lesotho (2016-2017) | 2435 | 15-59 | TND (1239), <LLOQ: 20 (402), <LLOQ: 400 (15) | 87.7 (86.1, 89.3) |
| Lesotho (2019-2020) | 3260 | 15+ | TND (1875), <LLOQ: 20 (406), <LLOQ: 40 (13), <LLOQ: 400 (10) | 91.5 (90.3, 92.6) |
| Malawi (2015-2016) | 1564 | 15-64 | TND (1156), <LLOQ: 40 (176) | 91.3 (89.3, 93.3) |
| Malawi (2020-2021) | 2167 | 15+ | TND (1712), <LLOQ: 40 (255), <LLOQ: 80 (43), <LLOQ: 839 (8) | 96.9 (96.0, 97.7) |
| Mozambique (2021-2022) | 1482 | 15+ | TND (622), <LLOQ: 20 (198), <LLOQ: 40 (14), <LLOQ: 400 (4) | 89.4 (87.3, 91.6) |
| Namibia (2017) | 2069 | 15-64 | TND (966), <LLOQ: 20 (343), <LLOQ: 40 (4), <LLOQ: 400 (5) | 91.3 (89.8, 92.9) |
| Nigeria (2018) | 1322 | 15-64 | TND (435), <LLOQ: 20 (250), <LLOQ: 40 (13), <LLOQ: 400 (5) | 80.9 (78.2, 83.6) |
| Rwanda (2018-2019) | 767 | 15-64 | TND (368), <LLOQ: 20 (165), <LLOQ: 40 (2) | 90.1 (87.6, 92.6) |
| Tanzania (2016-2017) | 1022 | 15+ | TND (477), <LLOQ: 20 (182), <LLOQ: 40 (3), <LLOQ: 400 (4) | 87.2 (84.5, 89.8) |
| Uganda (2016-2017) | 1185 | 15-64 | TND (470), <LLOQ: 20 (160) | 83.7 (81.4, 85.9) |
| Zambia (2016) | 1556 | 15-59 | TND (827), <LLOQ: 20 (281), <LLOQ: 550 (1) | 89.2 (87.4, 91.1) |
| Zambia (2021) | 1906 | 15+ | TND (1244), <LLOQ: 20 (254), <LLOQ: 400 (4) | 96.3 (95.0, 97.5) |
| Zimbabwe (2015-2016) | 2497 | 15+ | TND (1354), <LLOQ: 20 (418), <LLOQ: 182 (2) | 85.4 (83.6, 87.2) |
| Zimbabwe (2019-2020) | 2547 | 15+ | TND (1599), <LLOQ: 20 (367), <LLOQ: 40 (15), <LLOQ: 400 (3) | 90.3 (89.0, 91.7) |

TND – Target not detected, <LLOQ: 20 - less than lower limit of quantification of 20, <LLOQ: 40 - less than lower limit of quantification of 40, <LLOQ: 80 - less than lower limit of quantification of 80, <LLOQ: 182 - less than lower limit of quantification of 182, <LLOQ: 400 - less than lower limit of quantification of 400, <LLOQ: 550 - less than lower limit of quantification of 550, <LLOQ: 839 - less than lower limit of quantification of 839.

**Table S2.** Number of countries reporting viral load suppression data to UNAIDS at different thresholds for 2015 to 2023.

| Year (number of reporting countries) | Threshold for reporting viral load suppression in copies/mL |  |  |  |
| --- | --- | --- | --- | --- |
|  | <50 | <200 | <400 | <1000 |
| 2015 (27) | 0 | 5 | 0 | 22 |
| 2016 (46) | 0 | 7 | 0 | 39 |
| 2017 (69) | 0 | 11 | 0 | 58 |
| 2018 (91) | 0 | 11 | 0 | 80 |
| 2019 (90) | 1 | 10 | 0 | 79 |
| 2020 (98) | 2 | 11 | 1 | 84 |
| 2021 (98) | 2 | 13 | 1 | 82 |
| 2022 (96) | 2 | 11 | 1 | 82 |
| 2023 (91) | 2 | 10 | 1 | 78 |

Source: UNAIDS Estimates 2024 Spectrum files (https://hivtools.unaids.org/spectrum-file-request/)

**Figure S1A.**
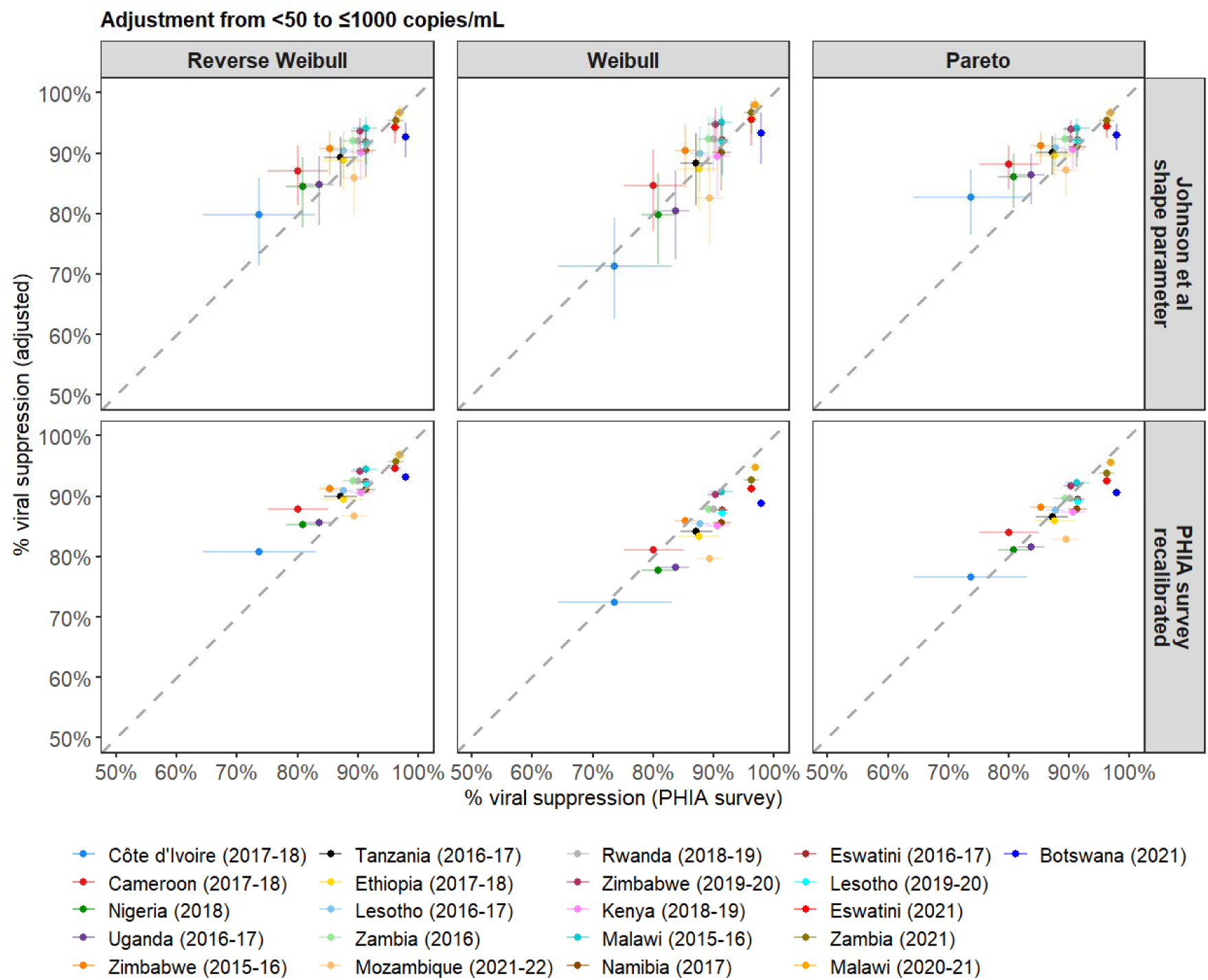
Scatter plots show the relationship between the observed percentage VLS estimates from the individual patient data in the PHIA surveys and the adjusted estimates (from <50 to ≤1000 copies/mL) using the reverse Weibull, Weibull and Pareto models and parameters from Johnson et al and calibration to PHIA survey data. Shape parameter for the Pareto model on calibration to PHIA survey data used in Figure is 1.20. Surveys in legend are sequenced in increasing order of observed VLS.

**Figure S1B.**
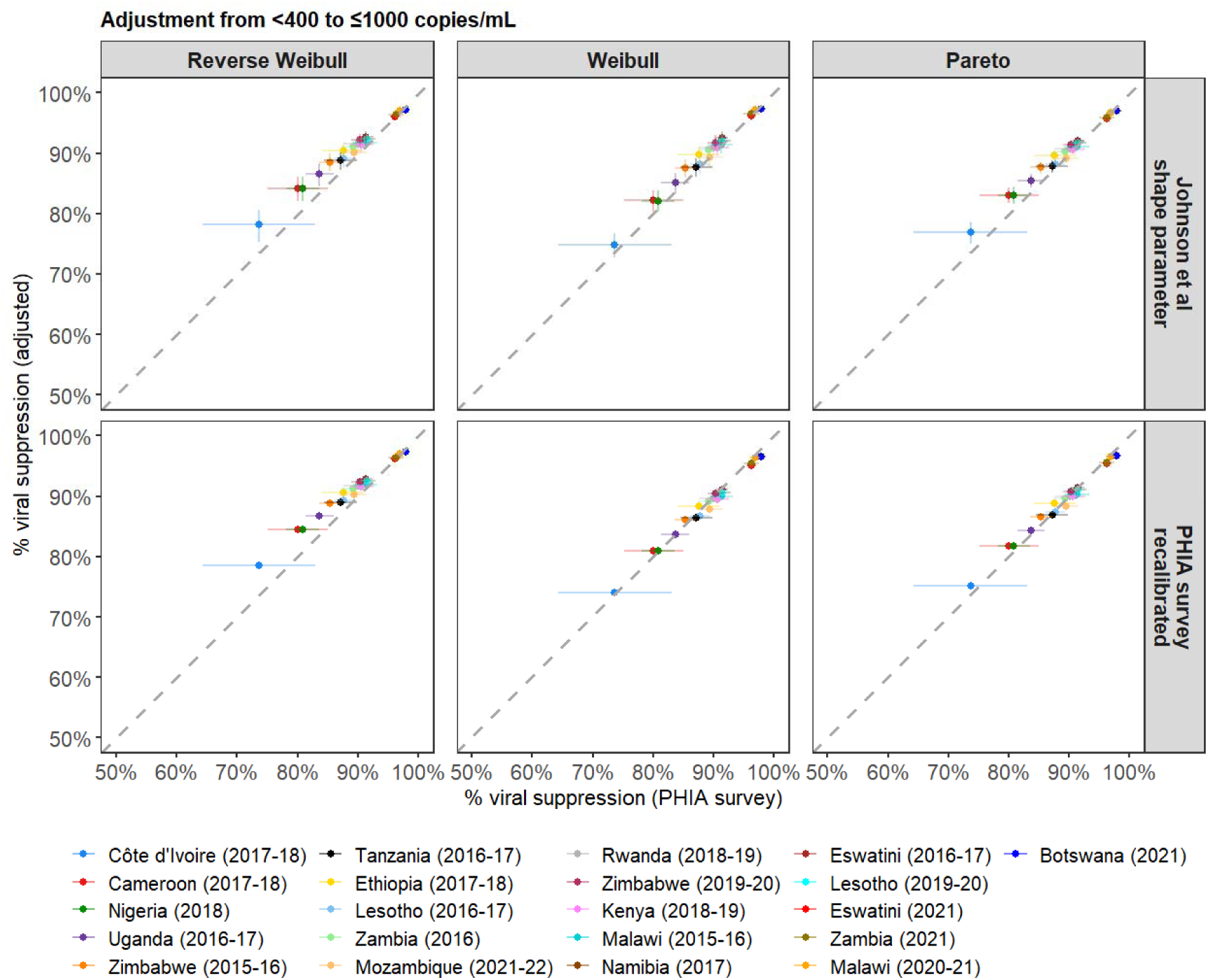
Scatter plots show the relationship between the observed percentage VLS estimates from the individual patient data in the PHIA surveys and the adjusted estimates (from <400 to ≤1000 copies/mL) using the reverse Weibull, Weibull and Pareto models and parameters from Johnson et al and calibration to PHIA survey data. Shape parameter for the Pareto model on calibration to PHIA survey data used in Figure is 1.20. Surveys in legend are sequenced in increasing order of observed VLS.

## Appendix S1. Modelling viral loads among PLHIV on ART using the Pareto distribution

The Pareto distribution was not suitable for likelihood-based fitting to individual-level survey observations because scale parameter *m* defines the lower limit of the distribution. However, we explored a range of plausible shape parameter values and assessed their visual fits to the PHIA data and how they performed in predicting the proportion with VL ≤1000 copies from the observed proportions <50, <200 or <400 copies/mL.

Plots of the empirical distribution of viral loads observed from the PHIA surveys and probability density estimates for the Pareto distribution using the shape parameter from Johnson *et al.* (shape = 1.73) showed that the Pareto model overestimates the probability of VLs being <1000 copies/mL in surveys with VLS <90% and underestimates this probability in surveys with VLS >90% **(Figure S2)**. A lower shape parameter value (shape = 1.20) improved the fit of the Pareto model to the empirical data in surveys with VLS <90% (**Figure S2A, S2B and S2C**) but increased the underestimation of the probability of VLs being <1000 copies/mL in surveys with VLS >90% **(Figure S2)**. Using higher shape parameter values (e.g. shape ≥ 2.00) compared to estimates from Johnson *et al.* improved the fits of the Pareto distribution to the observed data in surveys with VLS >90% but increased the overestimation of the probability of VLs being ≤1000 copies/mL in surveys with VLS <90% **(Figure S2)**.

The modelled lower limit of the Pareto distribution, which is scale parameter dependent, was in some cases higher than the lower bound of VL values among PLHIV on ART in the PHIA survey data. For example, when using the Pareto distribution and shape parameter from Johnson *et al.* to model 74% of VL values being ≤1000 copies/mL (observed in the Cote d’Ivoire, 2017-2018 survey), the scale parameter required translates to a lower bound VL value of 24 copies/mL, which was greater than the lower limit of detection (20 copies/mL) in the survey. To model lower levels of viral load suppression using the Pareto distribution required even larger scale parameters which translate to even greater lower bound values which exceed the lower limit observed in the empirical data **(Figure S4)**.

Figure S4 shows that for different shape parameter values for the Pareto distribution, the modelled VLS (VL ≤1000 copies/mL) at which the lower bound of the Pareto distribution would exceed the lower limit of VL (20 copies/mL) in the empirical data varies. For shape parameter values of 1.20, 1.73, 2.00 and 2.50, modelling ≤63%, ≤76%, ≤81% and ≤88% of VL values being ≤1000 copies/mL results in a lower bound VL value of ≥20 copies/mL **(Figure S4)**.

**Figure S2.**
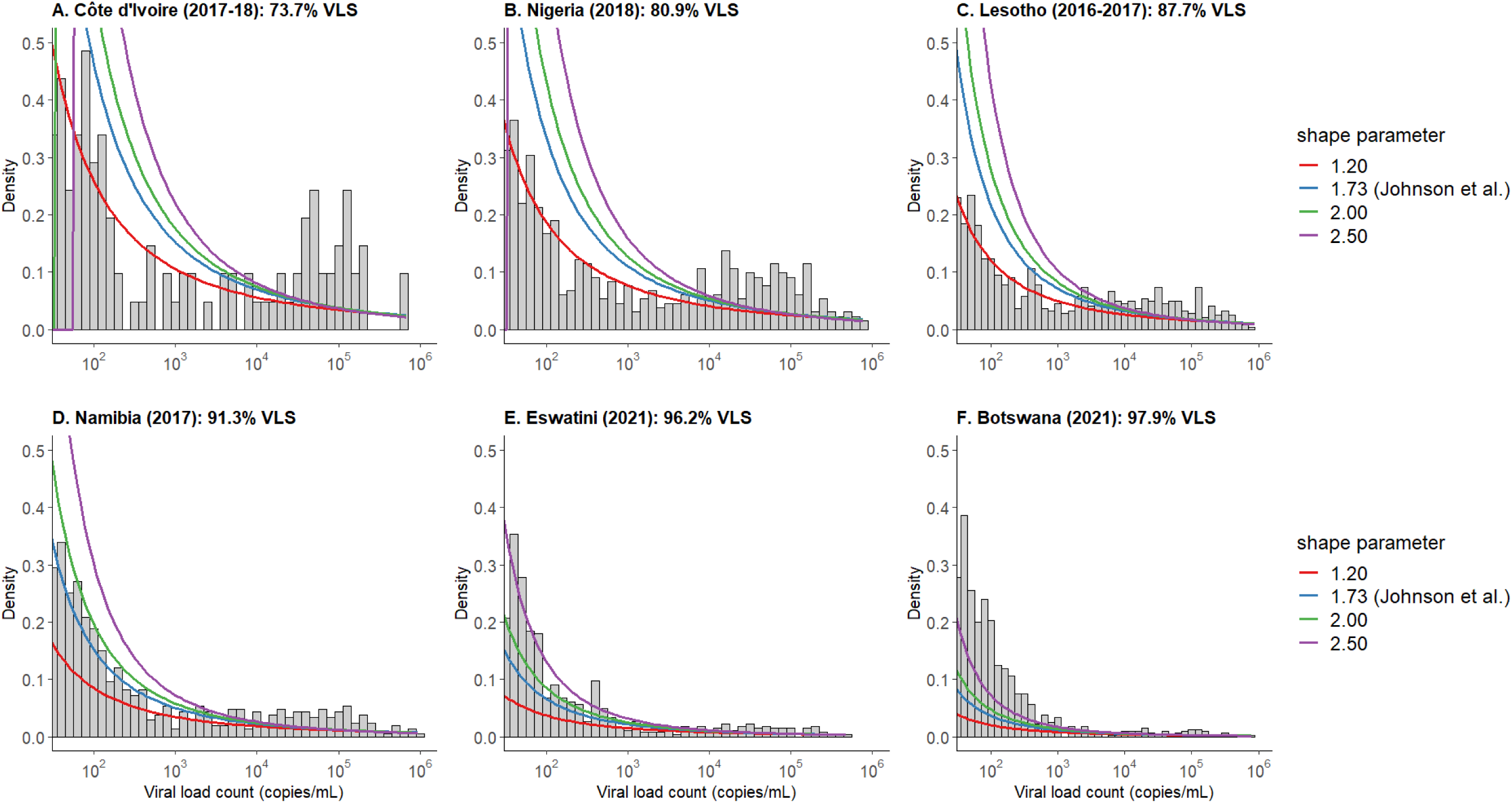
Histograms show the distribution of observed viral loads among PLHIV on ART in the A. Côte d’Ivoire (2017-18); B. Nigeria (2018); C. Lesotho (2016-2017); D. Namibia (2018); E. Eswatini (2021) and F. Botswana (2021) PHIA surveys. Lines show the probability density estimates for the Pareto distribution using shape parameters from Johnson *et al* (dashed line) and other plausible values. Note: the scale parameters were set so the cumulative probability of a viral load ≤1000 copies/mL is the same as VLS estimated from the survey data. The x-axis was truncated at 50 copies/mL for visualization purposes.

**Figure S3A.**
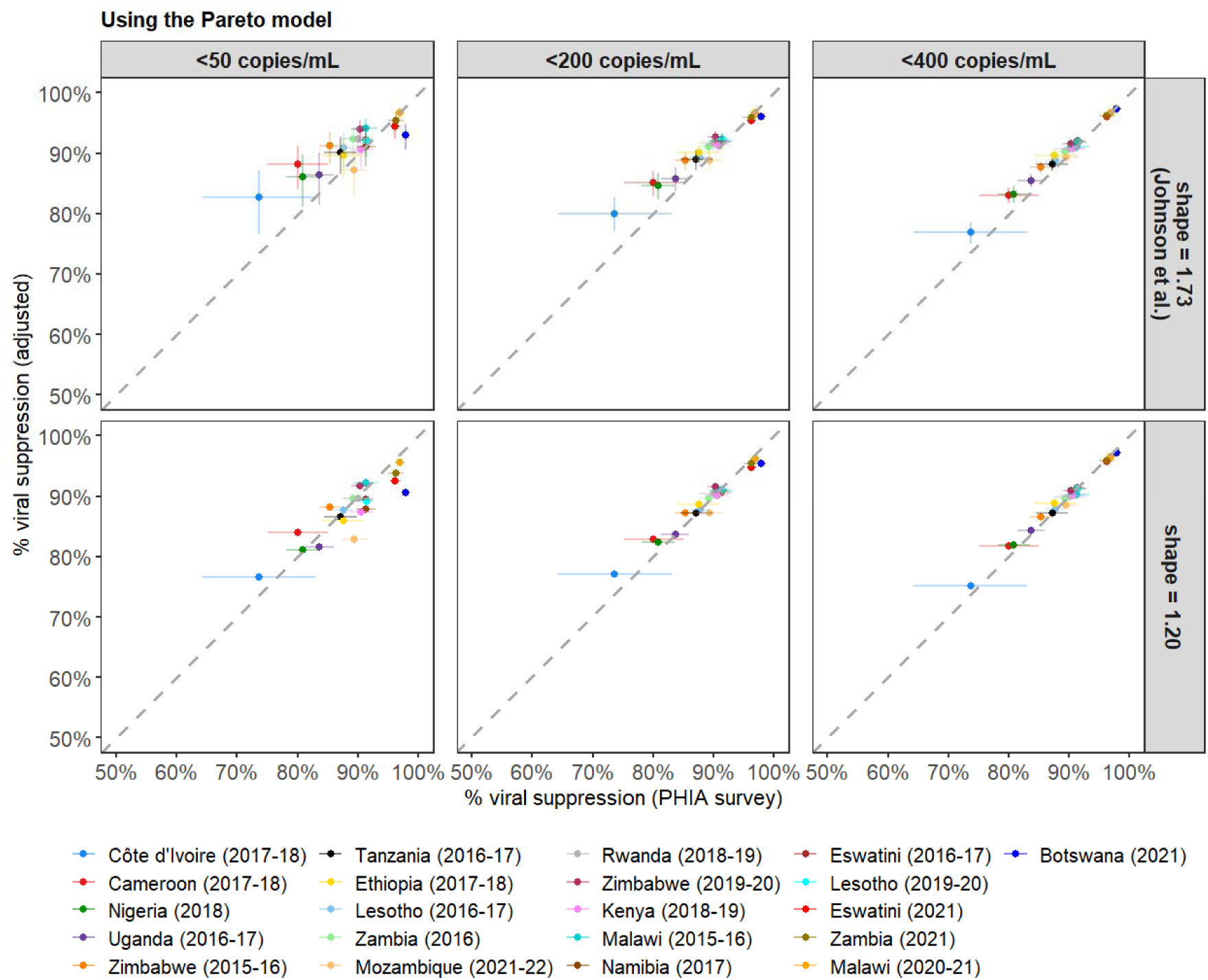
Scatter plots show the relationship between the observed percentage VLS estimates from the individual patient data in the PHIA surveys and the adjusted estimates (from <50, <200 and <400 to ≤1000 copies/mL) using the Pareto model and parameters from Johnson et al (shape = 1.73) or shape = 1.20. Root-mean-squared error (RMSE) when using the Pareto model with shape parameter 1.73 vs. 1.20 (RMSE for adjustment from <50 to ≤1000: 3.8% vs. 3.0%; RMSE for <200 to ≤1000: 2.5% vs. 1.4% and RMSE for <400 to ≤1000: 1.4% vs. 0.8%). Surveys in legend are sequenced in increasing order of observed VLS.

**Figure S3B.**
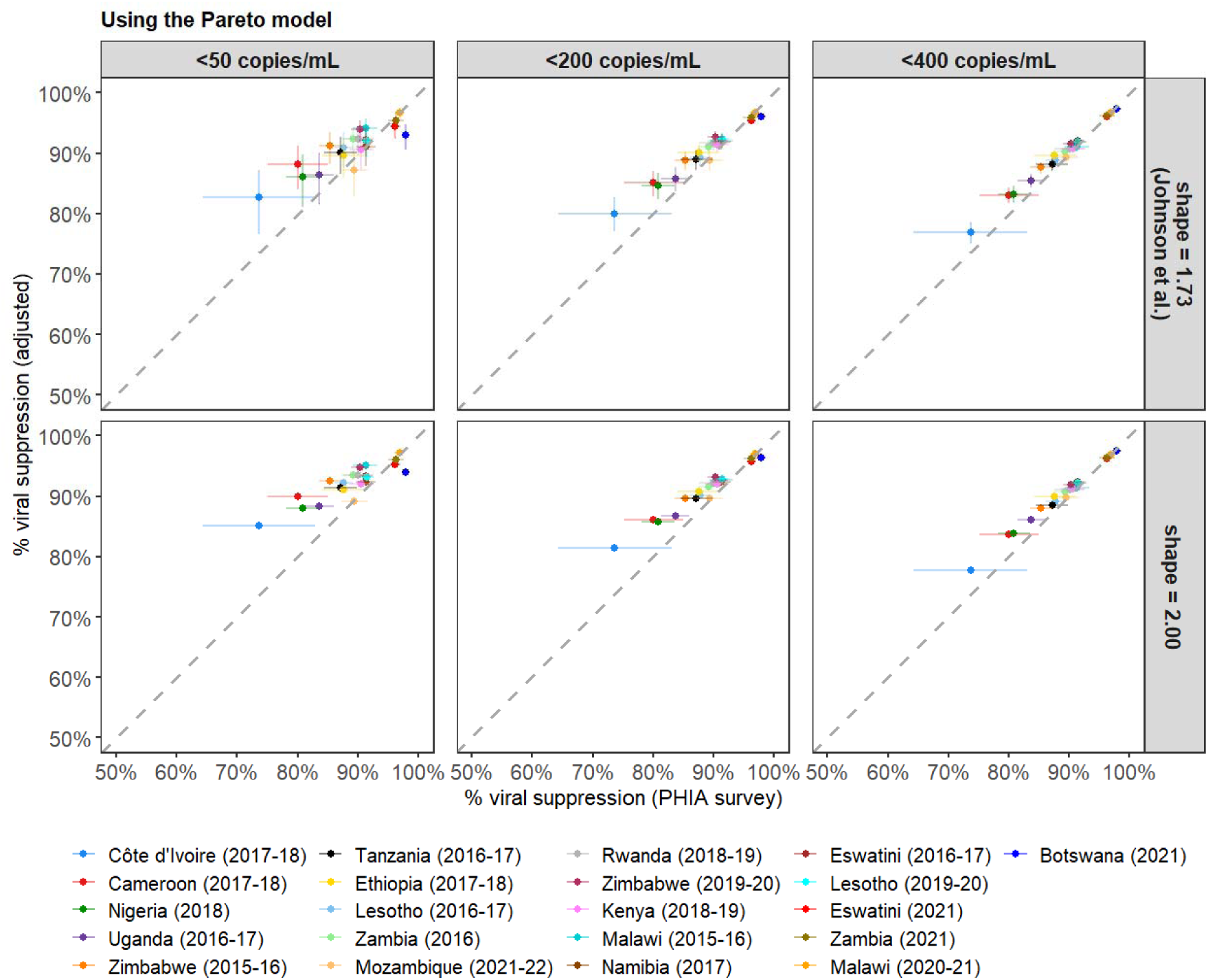
Scatter plots show the relationship between the observed percentage VLS estimates from the individual patient data in the PHIA surveys and the adjusted estimates (from <50, <200 and <400 to ≤1000 copies/mL) using the Pareto model and parameters from Johnson et al (shape = 1.73) or shape = 2.00. Root-mean-squared error (RMSE) when using the Pareto model with shape parameter 1.73 vs. 2.00 (RMSE for adjustment from <50 to ≤1000: 3.8% vs. 4.8%; RMSE for <200 to ≤1000: 2.5% vs. 3.1% and RMSE for <400 to ≤1000: 1.4% vs. 1.8%). Surveys in legend are sequenced in increasing order of observed VLS.

**Figure S3C.**
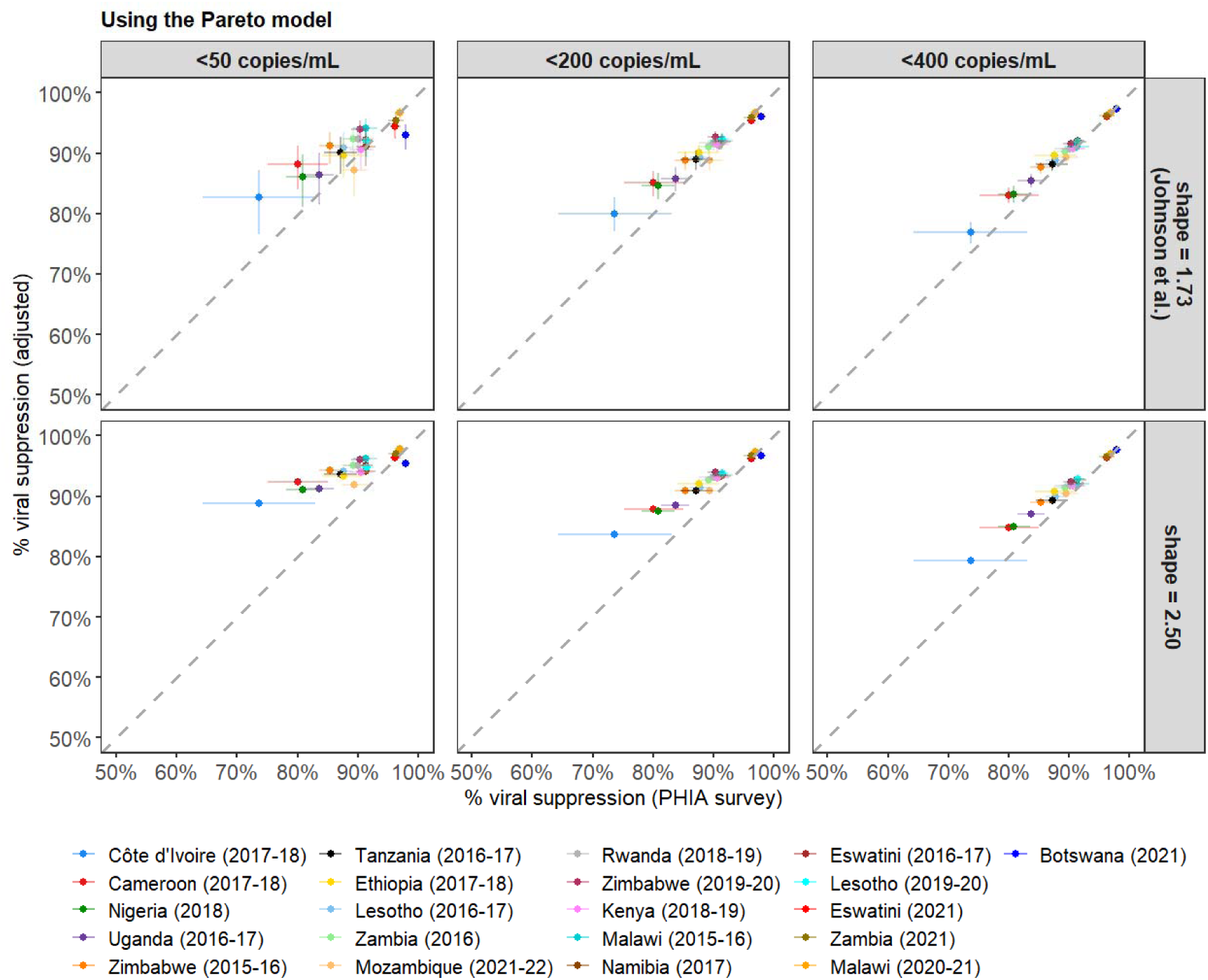
Scatter plots show the relationship between the observed percentage VLS estimates from the individual patient data in the PHIA surveys and the adjusted estimates (from <50, <200 and <400 to ≤1000 copies/mL) using the Pareto model and parameters from Johnson et al (shape = 1.73) or shape = 2.50. Root-mean-squared error (RMSE) when using the Pareto model with shape parameter 1.73 vs. 2.50 (RMSE for adjustment from <50 to ≤1000: 3.8% vs. 6.6%; RMSE for <200 to ≤1000: 2.5% vs. 4.2% and RMSE for <400 to ≤1000: 1.4% vs. 2.5%). Surveys in legend are sequenced in increasing order of observed VLS.

**Figure S4.**
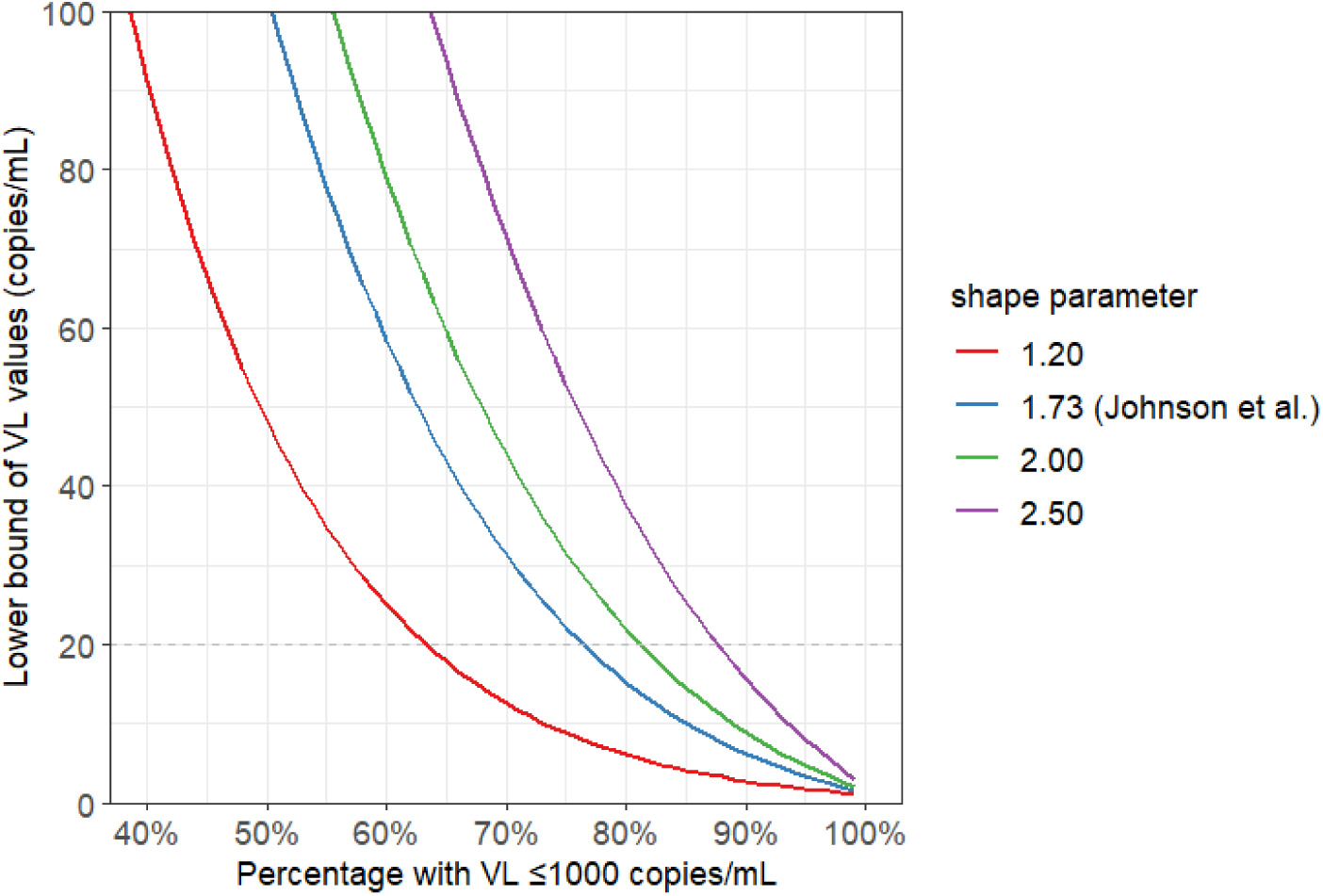
Plot shows the relationship between the lower bound (10^*m*^), where *m* is the scale parameter, and the proportion ≤1000 copies/mL for the Pareto distribution using different shape parameter values.

**Figure S5A.**
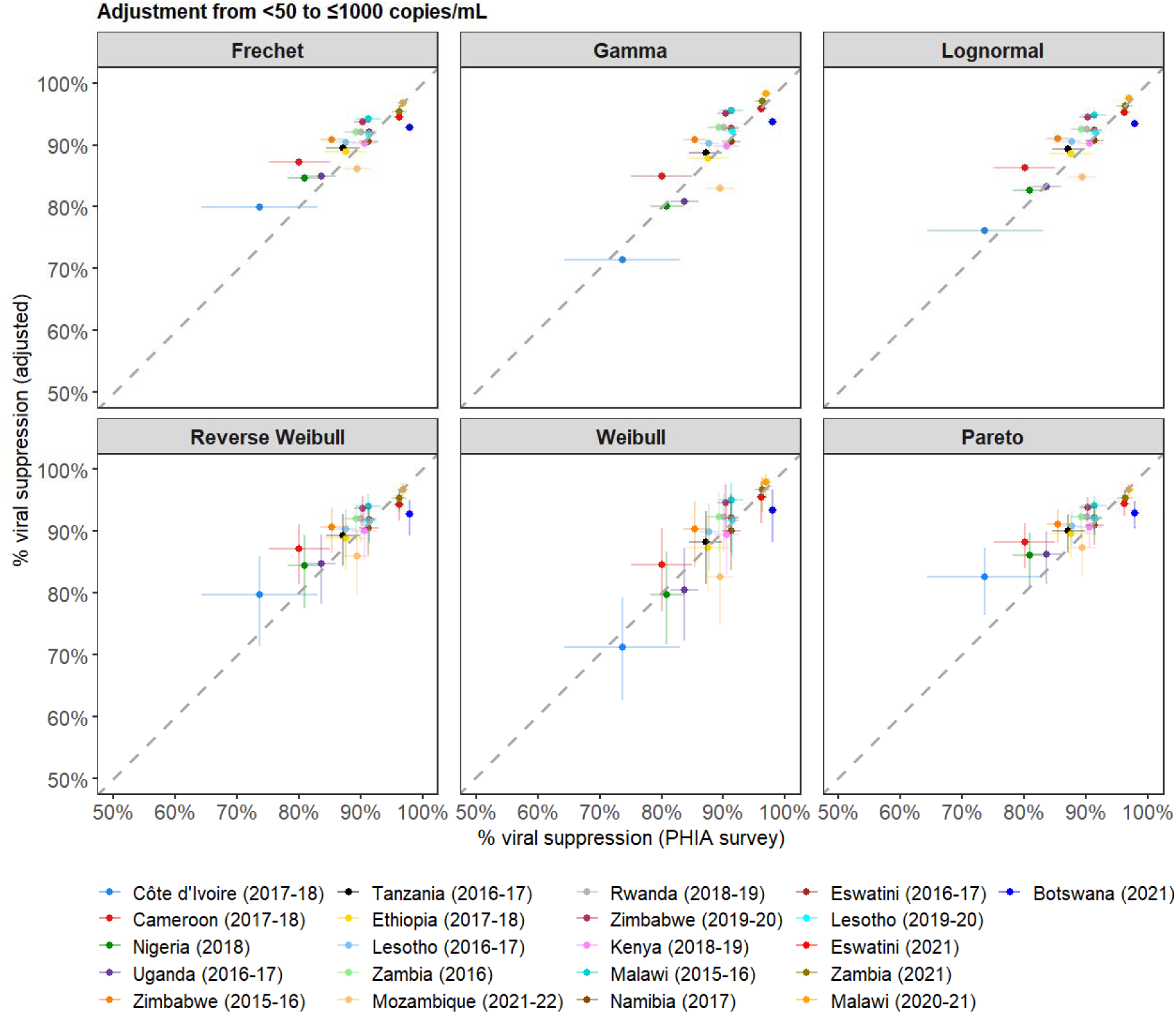
Scatter plots show the relationship between the observed percentage VLS estimates from the individual patient data in the PHIA surveys and the adjusted estimates (from <50 to ≤1000 copies/mL) using the Fréchet, gamma and lognormal model (parameters from calibration to the PHIA surveys), and the reverse Weibull, Weibull and Pareto models (using parameters from Johnson *et al*.). Surveys in legend are sequenced in increasing order of observed VLS.

**Figure S5B.**
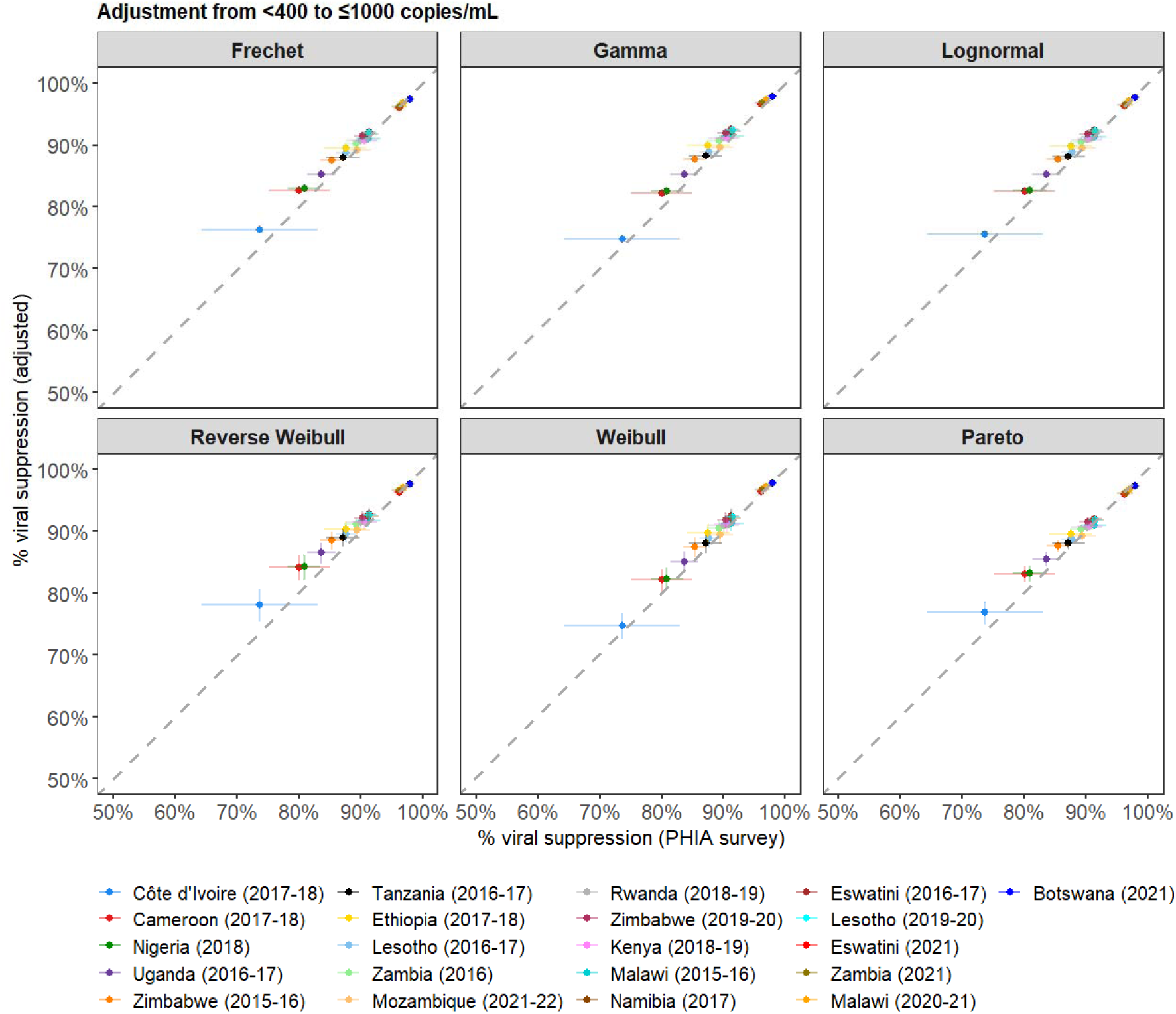
Scatter plots show the relationship between the observed percentage VLS estimates from the individual patient data in the PHIA surveys and the adjusted estimates (from <400 to ≤1000 copies/mL) using the Fréchet, gamma and lognormal model (parameters from calibration to the PHIA surveys), and the reverse Weibull, Weibull and Pareto models (using parameters from Johnson *et al*.). Surveys in legend are sequenced in increasing order of observed VLS.

**Figure S6.**
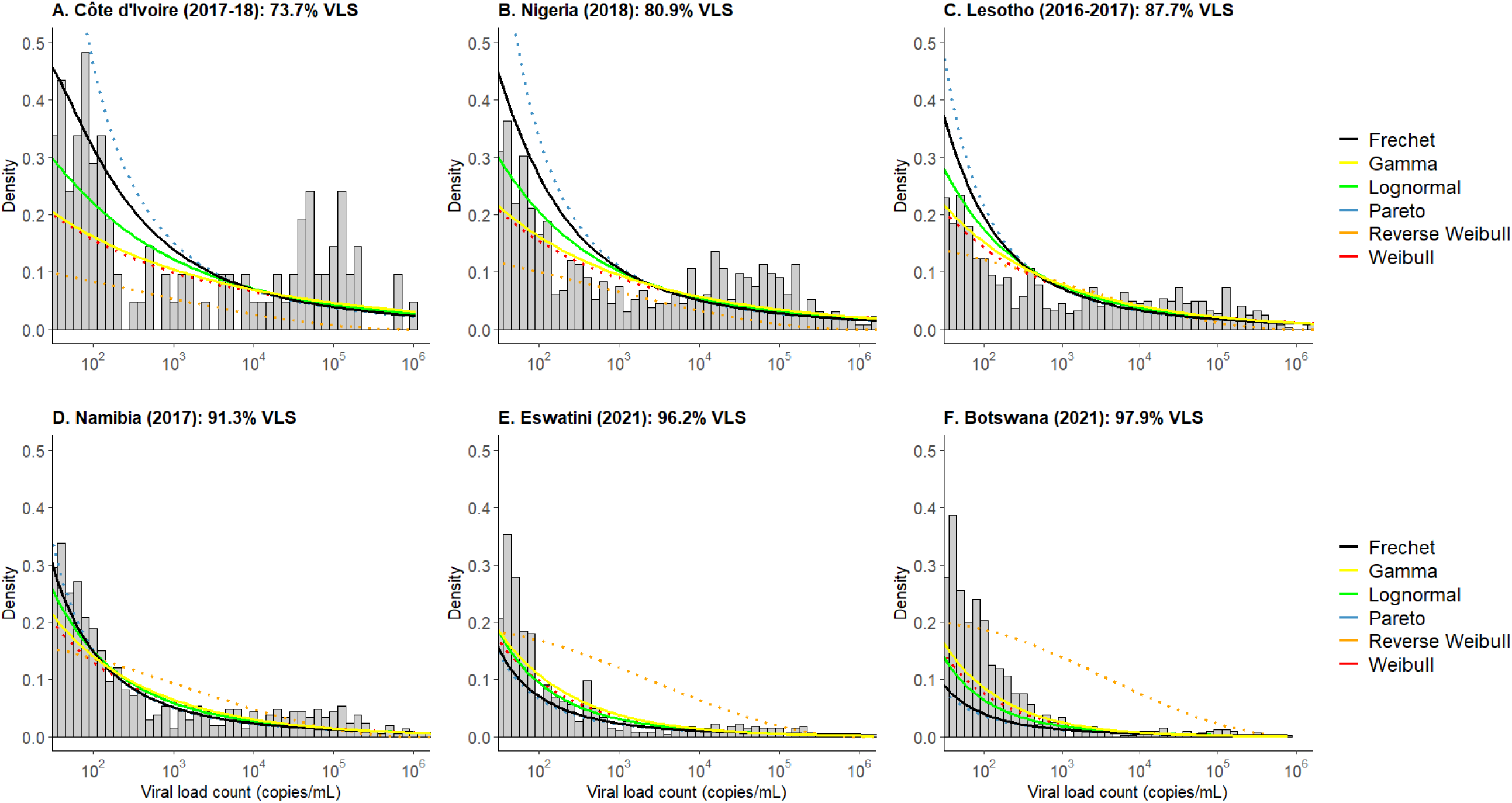
Histograms show the distribution of observed viral loads among PLHIV on ART in the A. Côte d’Ivoire (2017-18); B. Nigeria (2018); C. Lesotho (2016-2017); D. Namibia (2018); E. Eswatini (2021) and F. Botswana (2021) PHIA surveys. Lines show the probability density estimates for the Fréchet, gamma, lognormal using shape parameters from calibration to PHIA surveys (solid lines), and Pareto, reverse Weibull and Weibull models using shape parameters from Johnson *et al* (dotted lines). Note: the scale parameters were set so the cumulative probability of a viral load ≤1000 copies/mL is the same as VLS estimated from the survey data. The x-axis was truncated at 50 copies/mL for visualization purposes.

**Figure S7A.**
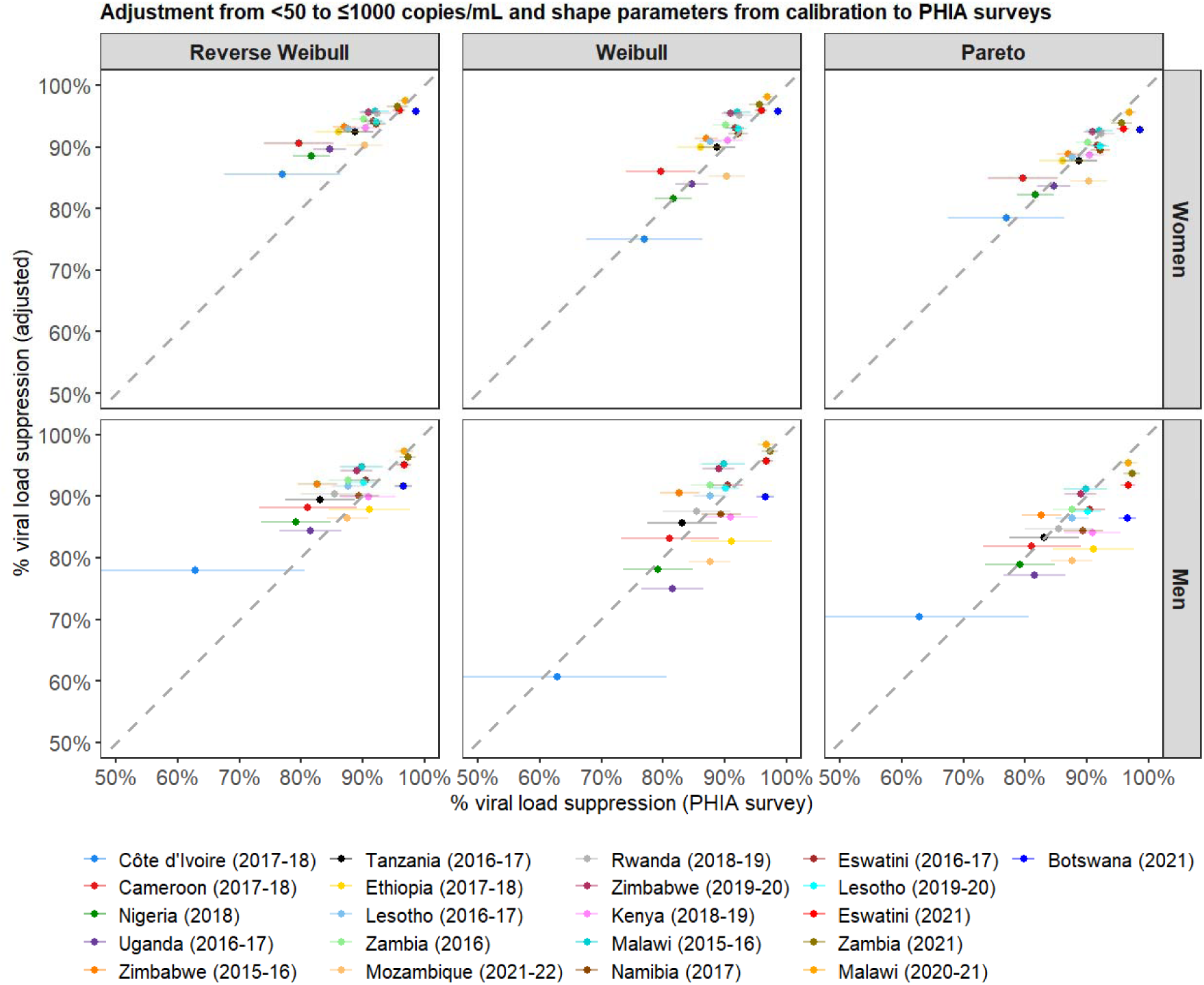
Scatter plots show the relationship between the observed percentage VLS estimates from the individual patient data in the PHIA surveys and the adjusted estimates (from <50 to ≤1000 copies/mL) by sex using the reverse Weibull, Weibull and Pareto models and parameters from Johnson *et al.* Surveys in legend are sequenced in increasing order of observed VLS.

**Figure 7B.**
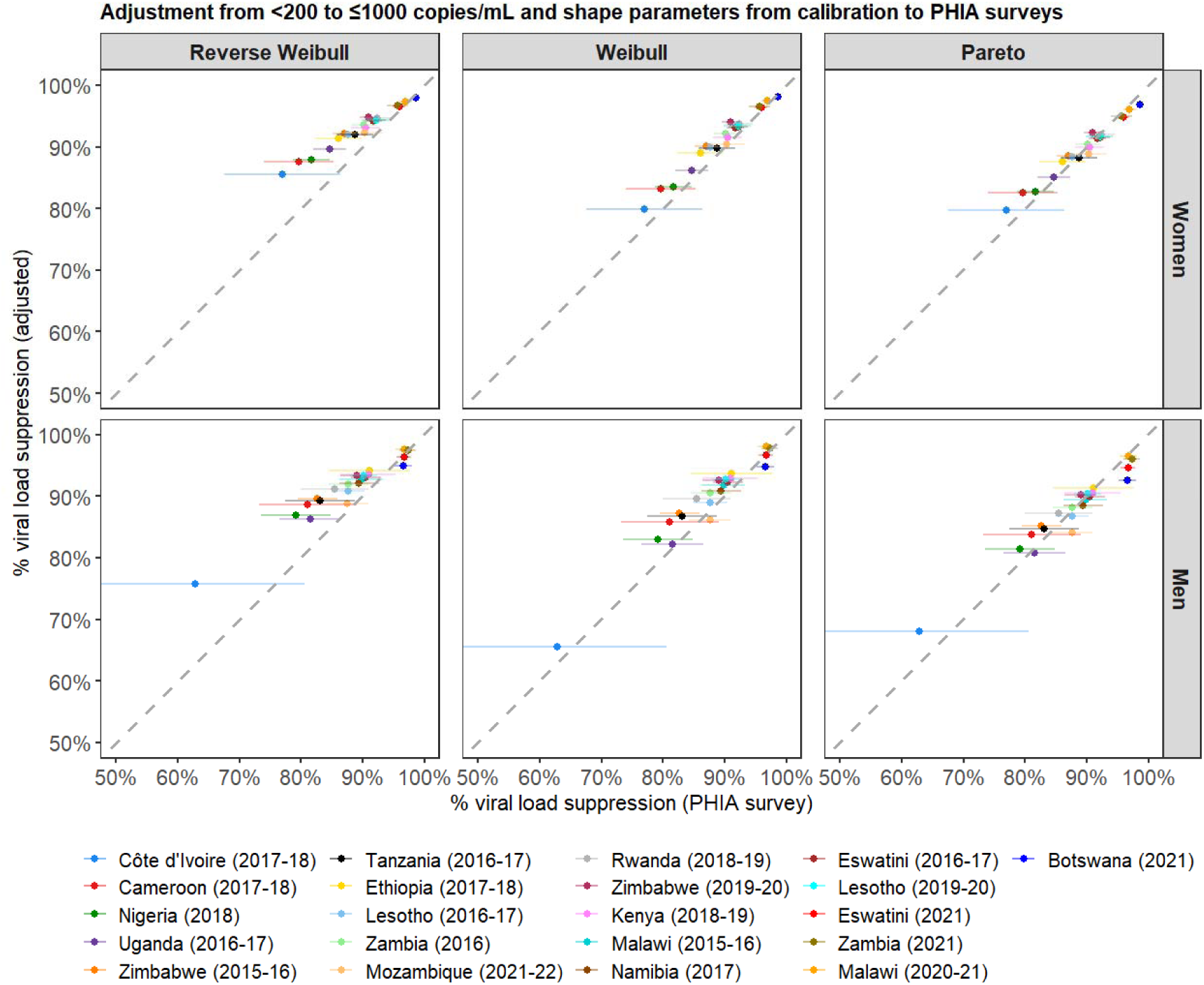
Scatter plots show the relationship between the observed percentage VLS estimates from the individual patient data in the PHIA surveys and the adjusted estimates (from <200 to ≤1000 copies/mL) by sex using the reverse Weibull, Weibull and Pareto models and parameters from Johnson *et al*. Surveys in legend are sequenced in increasing order of observed VLS

**Figure S7C.**
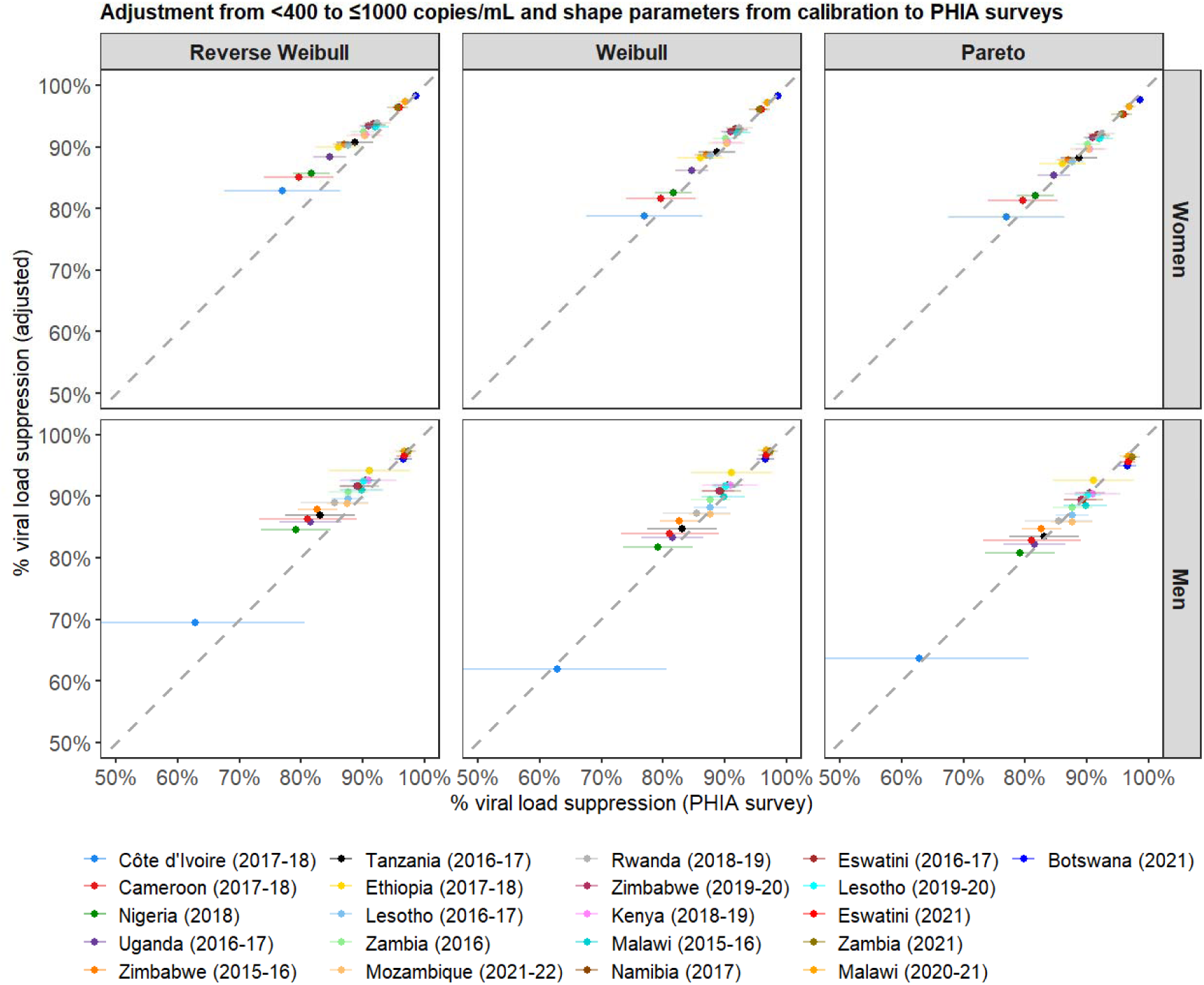
Scatter plots show the relationship between the observed percentage VLS estimates from the individual patient data in the PHIA surveys and the adjusted estimates (from <400 to ≤1000 copies/mL) by sex using the reverse Weibull, Weibull and Pareto models and parameters from Johnson *et al.* Surveys in legend are sequenced in increasing order of observed VLS.

**Figure S8A.**
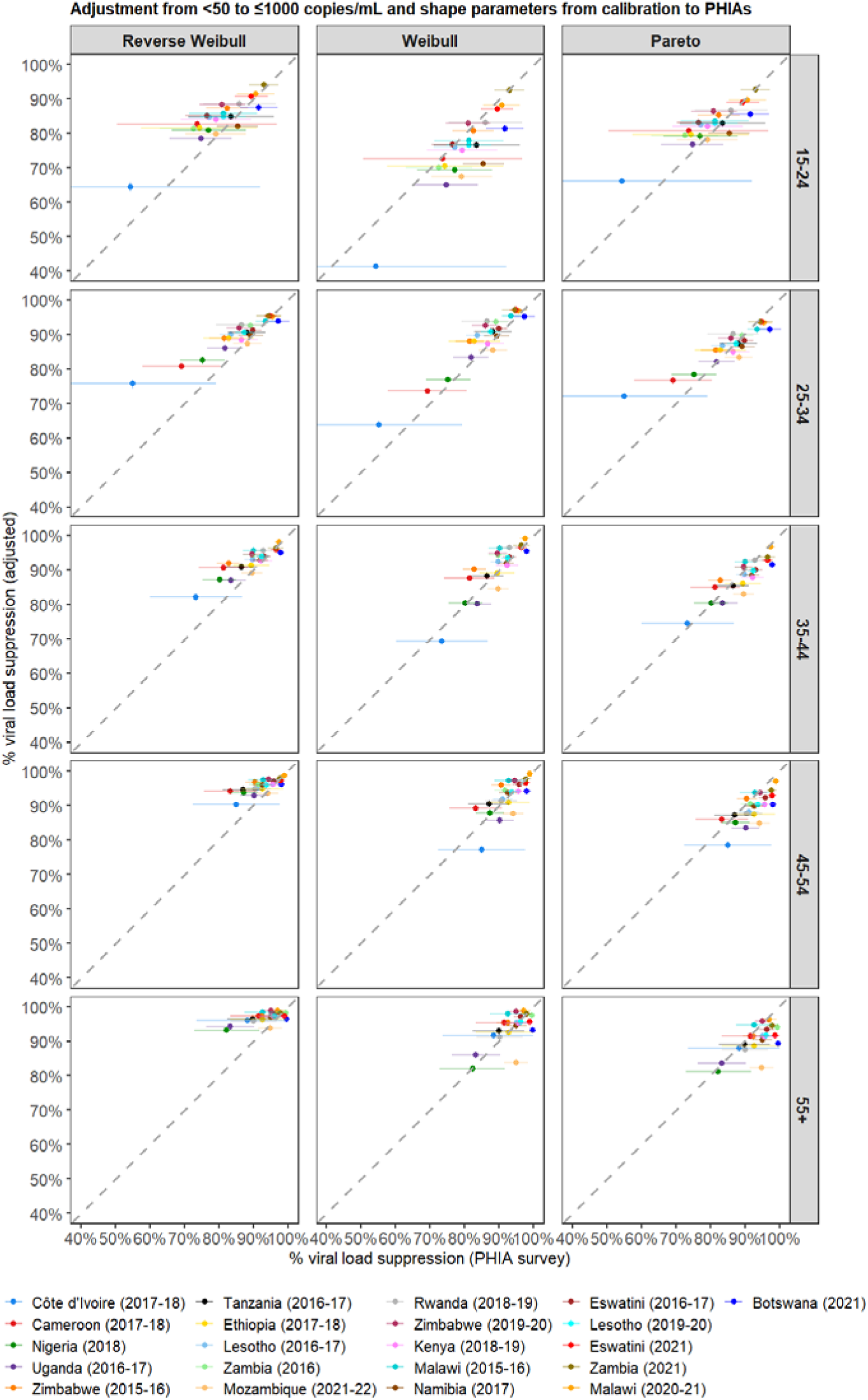
Scatter plots show the relationship between the observed percentage VLS estimates from the individual patient data in the PHIA surveys and the adjusted estimates (from <50 to ≤1000 copies/mL) by age using the reverse Weibull, Weibull and Pareto models and parameters from Johnson *et al.* Surveys in legend are sequenced in increasing order of observed VLS.

**Figure S8B.**
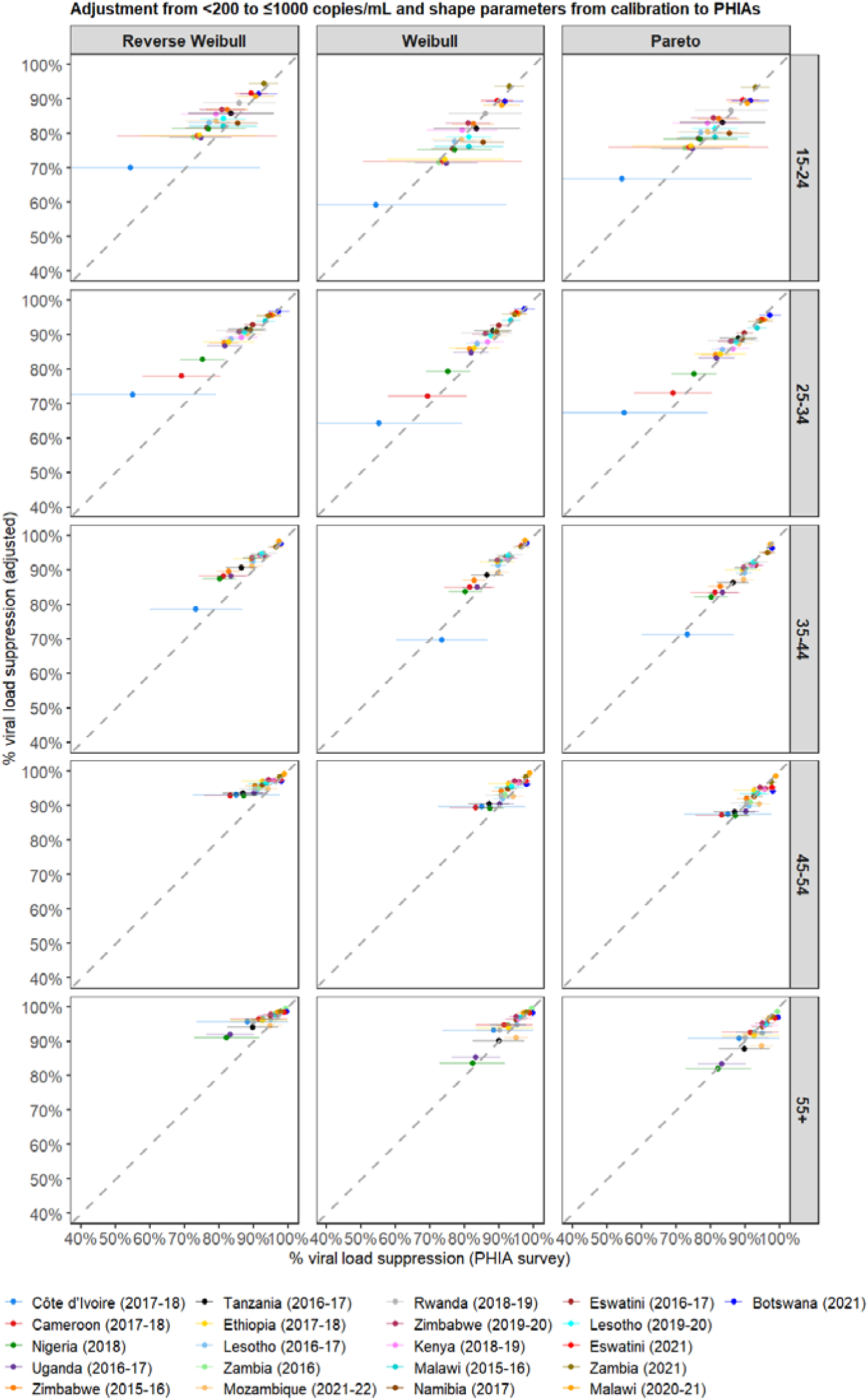
Scatter plots show the relationship between the observed percentage VLS estimates from the individual patient data in the PHIA surveys and the adjusted estimates (from <200 to ≤1000 copies/mL) by age using the reverse Weibull, Weibull and Pareto models and parameters from Johnson *et al.* Surveys in legend are sequenced in increasing order of observed VLS.

**Figure S8C.**
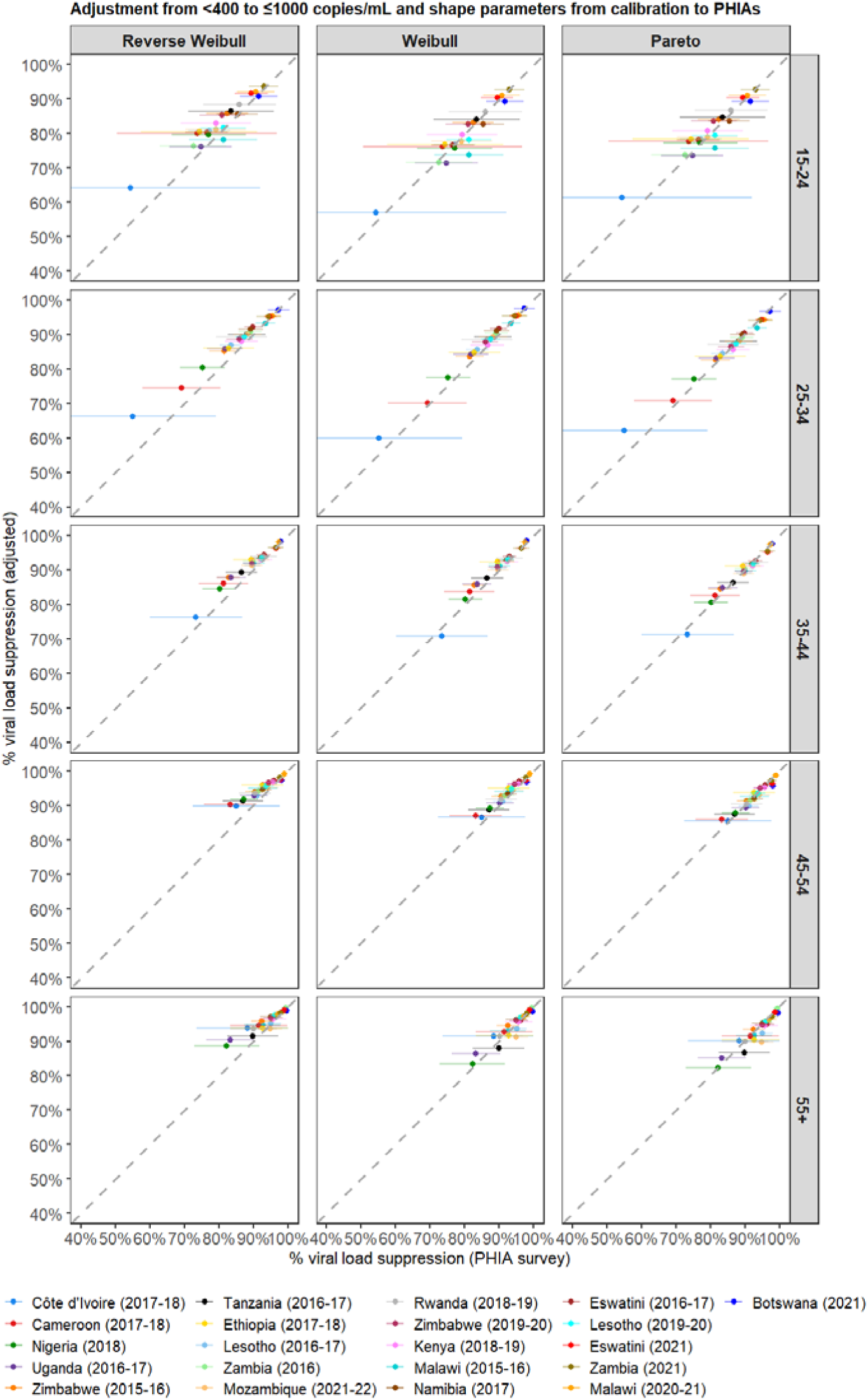
Scatter plots show the relationship between the observed percentage VLS estimates from the individual patient data in the PHIA surveys and the adjusted estimates (from <400 to ≤1000 copies/mL) by age using the reverse Weibull, Weibull and Pareto models and parameters from Johnson *et al.* Surveys in legend are sequenced in increasing order of observed VLS.

**Table S3.**
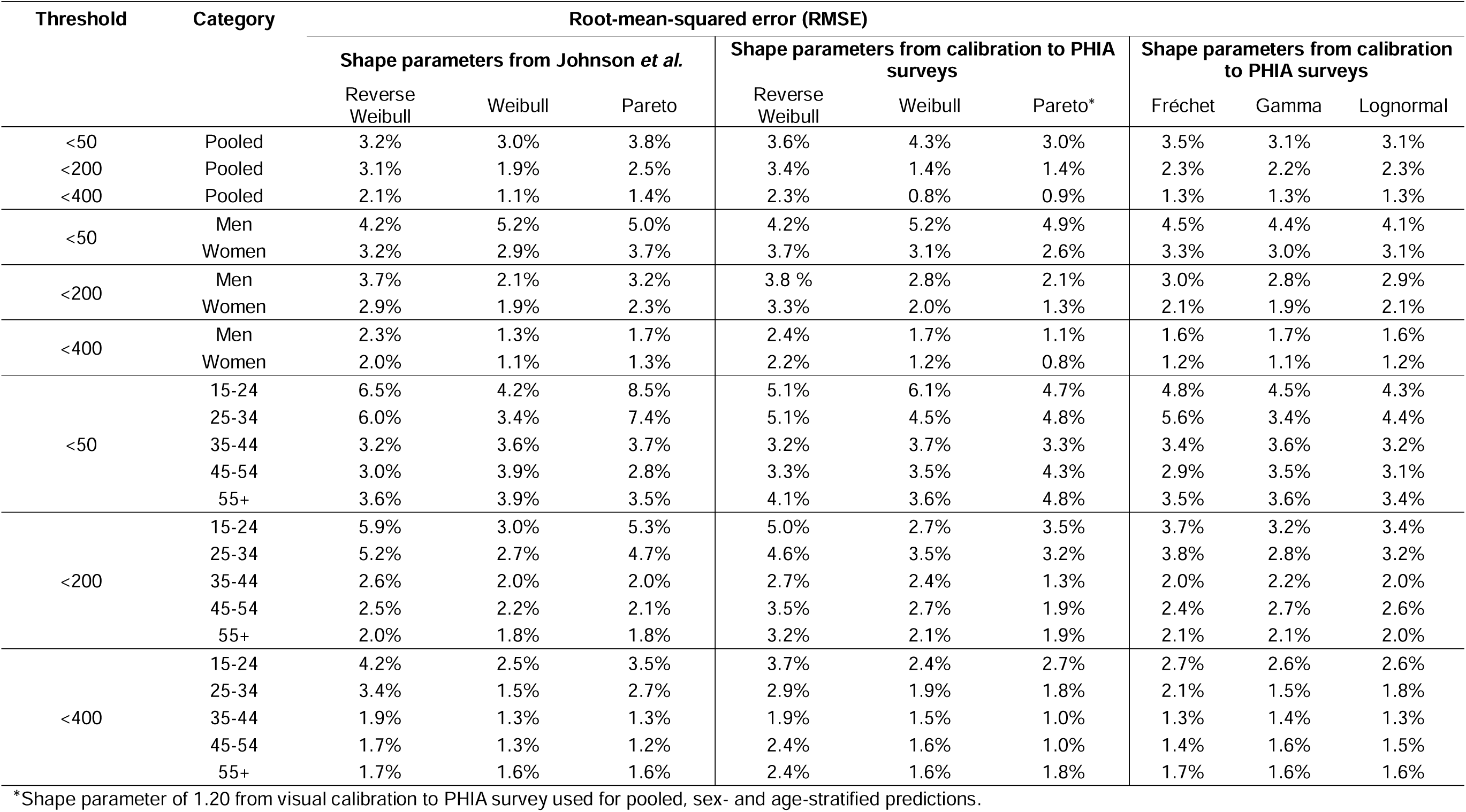
Root-mean-squared error (which quantifies the square root of the variance of residuals between the predicted values from the model and actual values from the PHIA survey data). A lower RMSE indicates better model fit and more precise model predictions. A shape parameter of 1.20 was used for the pooled, sex- and age-stratified comparisons for the Pareto model.

**Table S4.** Average bias (which quantifies the average difference between the models’ predicted VLS point estimate, and the actual survey point estimate). Note: negative values indicate underestimation while positive values indicate overestimation.

| Threshold | Category | Average bias |  |  |  |  |  |  |  |  |
| --- | --- | --- | --- | --- | --- | --- | --- | --- | --- | --- |
|  |  | Johnson *et al.* |  |  | Calibration to PHIA surveys |  |  | Calibration to PHIA surveys |  |  |
|  |  | Reverse Weibull | Weibull | Pareto | Reverse Weibull | Weibull | Pareto | Fréchet | Gamma | Lognormal |
| <50 | Pooled | 1.3% | 0.4% | 2.0% | 1.9% | -3.4% | 1.2% | 1.7% | 1.0% | 1.6% |
| <200 | Pooled | 2.3% | 1.5% | 1.5% | 2.6% | -0.8% | 0.1% | 1.5 % | 1.8% | 1.7% |
| <400 | Pooled | 1.6% | 0.8% | 0.8% | 1.8% | -0.4% | 0.2% | 0.8% | 1.0% | 0.9% |
| <50 | Men | 0.5% | -2.1% | 1.6% | 0.7% | 0.2% | -2.2% | 1.4% | 0.2% | 0.9% |
|  | Women | 1.7% | 1.3% | 2.2% | 2.3% | 1.6% | -0.7% | 1.9% | 1.4% | 1.9% |
| <200 | Men | 2.5% | 1.2% | 1.8% | 2.6% | 2.2% | 0.1% | 1.8% | 2.2% | 2.1% |
|  | Women | 2.2% | 1.5% | 1.5% | 2.6% | 1.7% | 0.1% | 1.4% | 1.6% | 1.6% |
| <400 | Men | 1.7% | 0.7% | 0.9% | 1.8% | 1.2% | 0.05% | 1.0% | 1.2% | 1.1% |
|  | Women | 1.8% | 0.7% | 0.9% | 1.7% | 1.0% | <0.0001% | 0.7% | 0.9% | 0.8% |
| <50 | 15-24 | 5.2% | 1.0% | 6.8% | 3.4% | -4.3% | 2.3% | 3.1% | 1.8% | 2.5% |
|  | 25-34 | 3.5% | 1.8% | 4.5% | 2.4% | 3.4% | 1.0% | 3.0% | 1.8% | 2.6% |
|  | 35-44 | 1.0% | 0.03% | 1.7% | 1.1% | 1.5% | -1.4% | 1.5% | 0.6% | 1.3% |
|  | 45-54 | -0.8% | -1.0% | -0.3% | 1.6% | 0.4% | -3.1% | 0.6% | 0.3% | 0.7% |
|  | 55+ | -1.1% | -1.0% | -0.7% | 2.0% | 0.5% | -3.2% | 0.7% | 0.5% | 0.7% |
| <200 | 15-24 | 4.6% | 1.5% | 3.7% | 3.6% | -0.9% | 1.3% | 1.9% | 1.9% | 1.9% |
|  | 25-34 | 3.7% | 2.1% | 2.9% | 3.1% | 2.9% | 1.1% | 2.2% | 2.1% | 2.2% |
|  | 35-44 | 1.9% | 1.0% | 1.2% | 2.0% | 1.7% | -0.2% | 1.2% | 1.3% | 1.3% |
|  | 45-54 | 1.4% | 1.2% | 0.7% | 2.5% | 1.8% | -0.3% | 1.3% | 1.8% | 1.6% |
|  | 55+ | 0.6% | 0.5% | 0.04% | 2.2% | 1.1% | -0.9% | 0.7% | 1.1% | 0.9% |
| <400 | 15-24 | 3.1% | 0.9% | 2.1% | 2.5% | -0.4% | 0.7% | 1.0% | 1.1% | 1.1% |
|  | 25-34 | 2.4% | 1.1% | 1.5% | 2.0% | 1.5% | 0.5% | 1.1% | 1.1% | 1.1% |
|  | 35-44 | 1.4% | 0.6% | 0.6% | 1.4% | 1.0% | -0.1% | 0.6% | 0.8% | 0.7% |
|  | 45-54 | 1.1% | 0.8% | 0.5% | 1.8% | 1.1% | -0.08% | 0.8% | 1.1% | 1.0% |
|  | 55+ | 0.3% | 0.02% | -0.3% | 1.3% | 0.4% | -0.8% | 0.1% | 0.4% | 0.3% |

**Table S5.** Comparing root-mean-squared error in adjustments from <50, <200 and <400 to ≤1000 copies/mL by sex and age using the Fréchet, gamma and lognormal models and common versus sex- and age-specific shape parameters from calibration to the PHIA survey data.

| Threshold | Category | Root-mean-squared error (RMSE) |  |  |  |  |  |
| --- | --- | --- | --- | --- | --- | --- | --- |
|  |  | Common shape parameter from calibration to PHIA survey data |  |  | Age- and sex-specific shape parameters from calibration to PHIA survey data |  |  |
|  |  | Fréchet | Gamma | Lognormal | Fréchet | Gamma | Lognormal |
| <50 | Men | 4.3% | 5.0% | 4.0% | 4.5% | 4.4% | 4.1% |
|  | Women | 3.5% | 3.2% | 3.3% | 3.3% | 3.0% | 3.1% |
| <200 | Men | 2.8% | 2.3% | 2.6% | 3.0% | 2.8% | 2.9% |
|  | Women | 2.2% | 2.1% | 2.2% | 2.1% | 1.9% | 2.1% |
| <400 | Men | 1.5% | 1.5% | 1.5% | 1.6% | 1.7% | 1.6% |
|  | Women | 1.3% | 1.2% | 1.3% | 1.2% | 1.1% | 1.2% |
| <50 | 15-24 | 7.1% | 4.5% | 5.4% | 4.8% | 4.5% | 4.3% |
|  | 25-34 | 6.4% | 3.7% | 5.1% | 5.6% | 3.4% | 4.4% |
|  | 35-44 | 3.4% | 3.6% | 3.2% | 3.4% | 3.6% | 3.2% |
|  | 45-54 | 2.9% | 3.7% | 3.1% | 2.9% | 3.5% | 3.1% |
|  | 55+ | 3.5% | 3.8% | 3.5% | 3.5% | 3.6% | 3.4% |
| <200 | 15-24 | 4.7% | 3.2% | 3.9% | 3.7% | 3.2% | 3.4% |
|  | 25-34 | 4.2% | 3.0% | 3.6% | 3.8% | 2.8% | 3.2% |
|  | 35-44 | 1.9% | 2.2% | 2.0% | 2.0% | 2.2% | 2.0% |
|  | 45-54 | 2.1% | 2.4% | 2.3% | 2.4% | 2.7% | 2.6% |
|  | 55+ | 1.8% | 1.9% | 1.8% | 2.1% | 2.1% | 2.0% |
| <400 | 15-24 | 3.2% | 2.6% | 2.8% | 2.7% | 2.6% | 2.6% |
|  | 25-34 | 2.4% | 1.6% | 2.0% | 2.1% | 1.5% | 1.8% |
|  | 35-44 | 1.3% | 1.4% | 1.3% | 1.3% | 1.4% | 1.3% |
|  | 45-54 | 1.2% | 1.4% | 1.3% | 1.4% | 1.6% | 1.5% |
|  | 55+ | 1.6% | 1.6% | 1.6% | 1.7% | 1.6% | 1.6% |

**Table S6.** Comparing average bias in adjustments from <50, <200 and <400 to ≤1000 copies/mL by sex and age using the Fréchet, gamma and lognormal models and common versus sex- and age-specific shape parameters from calibration to the PHIA survey data.

| Threshold | Category | Average bias |  |  |  |  |  |
| --- | --- | --- | --- | --- | --- | --- | --- |
|  |  | Common shape parameter from calibration to PHIA survey data |  |  | Age- and sex-specific shape parameters from calibration to PHIA survey data |  |  |
|  |  | Fréchet | Gamma | Lognormal | Fréchet | Gamma | Lognormal |
| <50 | Men | 1.0% | -1.4% | 0.06% | 1.4% | 0.2% | 0.9% |
|  | Women | 2.0% | 1.9% | 2.2% | 1.9% | 1.4% | 1.9% |
| <200 | Men | 1.6% | 1.6% | 1.7% | 1.8% | 2.2% | 2.1% |
|  | Women | 1.5% | 1.8% | 1.7% | 1.4% | 1.6% | 1.6% |
| <400 | Men | 0.9% | 0.9% | 0.9% | 1.0% | 1.2% | 1.1% |
|  | Women | 0.7% | 1.0% | 0.9% | 0.7% | 0.9% | 0.8% |
| <50 | 15-24 | 5.7% | 1.7% | 4.0% | 3.1% | 1.8% | 2.5% |
|  | 25-34 | 4.0% | 2.4% | 3.5% | 3.0% | 1.8% | 2.6% |
|  | 35-44 | 1.4% | 0.07% | 1.3% | 1.5% | 0.6% | 1.3% |
|  | 45-54 | -0.4% | -0.5% | -0.3% | 0.6% | 0.3% | 0.7% |
|  | 55+ | -0.7% | -0.4% | -0.4% | 0.7% | 0.5% | 0.7% |
| <200 | 15-24 | 3.3% | 1.9% | 2.7% | 1.9% | 1.9% | 1.9% |
|  | 25-34 | 2.7% | 2.4% | 2.6% | 2.2% | 2.1% | 2.2% |
|  | 35-44 | 1.1% | 1.3% | 1.3% | 1.2% | 1.3% | 1.3% |
|  | 45-54 | 0.8% | 1.5% | 1.2% | 1.3% | 1.8% | 1.6% |
|  | 55+ | 0.08% | 0.7% | 0.5% | 0.7% | 1.1% | 0.9% |
| <400 | 15-24 | 1.8% | 1.1% | 1.5% | 1.0% | 1.1% | 1.1% |
|  | 25-34 | 1.4% | 1.3% | 1.4% | 1.1% | 1.1% | 1.1% |
|  | 35-44 | 0.6% | 0.8% | 0.7% | 0.6% | 0.8% | 0.7% |
|  | 45-54 | 0.5% | 0.9% | 0.8% | 0.8% | 1.1% | 1.0% |
|  | 55+ | -0.2% | 0.2% | -0.00004% | 0.1% | 0.4% | 0.3% |

## References

1. World Health Organization. What’s new in treatment monitoring: viral load and CD4 testing [Internet]. 2017 [cited 2021 Nov 27]. Available from: https://www.who.int/publications/i/item/WHO-HIV-2017.22

2. Joint United Nations Programme on HIV/AIDS (UNAIDS). 90-90-90 An ambitious treatment target to help end the AIDS epidemic [Internet]. Vol. 2021. 2014 [cited 2024 Nov 1]. Available from: https://www.unaids.org/en/resources/documents/2017/90-90-90

3. Joint United Nations Programme on HIV/AIDS (UNAIDS). Prevailing against pandemics by putting people at the centre — World AIDS Day report 2020 [Internet]. 2020 [cited 2024 Mar 25]. Available from: https://aidstargets2025.unaids.org/assets/images/prevailing-against-pandemics_en.pdf

4. Raymond A, Hill A, Pozniak A. Large disparities in HIV treatment cascades between eight European and high-income countries – analysis of break points. J Int AIDS Soc. 2014;17(4S3).

5. Drew RS, Rice B, Rüütel K, Delpech V, Attawell KA, Hales DK, et al. HIV continuum of care in Europe and Central Asia. HIV Med. 2017;18(7).

6. Gourlay AJ, Pharris AM, Noori T, Supervie V, Rosinska M, Van Sighem A, et al. Towards standardized definitions for monitoring the continuum of HIV care in Europe. AIDS. 2017;31(15).

7. Erly S, Campos L, Buskin S, Reuer J. Evaluating surveillance definitions of HIV viral suppression 2015–2019: Which definition best detected barriers to care? J Public Health Res. 2023;12(2).

8. Johnson LF, Kariminia A, Trickey A, Yiannoutsos CT, Ekouevi DK, Minga AK, et al. Achieving consistency in measures of HIV-1 viral suppression across countries: derivation of an adjustment based on international antiretroviral treatment cohort data. J Int AIDS Soc. 2021;24(S5).

9. Song JW, Yang G, Kamara MN, Sun W, Guan Q, Barrie U, et al. HIV viral suppression at different thresholds and duration of treatment in the dolutegravir treatment era in Sierra Leone: a nationwide survey. Virol J. 2023;20(1).

10. Chun HM, Milligan K, Boyd MA, Abutu A, Bachanas P, Dirlikov E. Reaching HIV epidemic control in Nigeria using a lower HIV viral load suppression cut-off. AIDS. 2023;37(13).

11. Rosen JG, Reynolds SJ, Galiwango RM, Kigozi G, Quinn TC, Ratmann O, et al. A moving target: impacts of lowering viral load suppression cutpoints on progress towards HIV epidemic control goals. Vol. 37, AIDS. 2023.

12. World Health Organization. WHO Technical Report: HIV Drug Resistance Report 2021 [Internet]. 2021 [cited 2024 May 13]. Available from: https://iris.who.int/bitstream/handle/10665/349340/9789240038608-eng.pdf

13. ICAP at Columbia University. PHIA Project: Population-based HIV Impact Assessment [Internet]. 2021 [cited 2025 Jan 11]. Available from: https://phia.icap.columbia.edu/

14. Sachathep K, Radin E, Hladik W, Hakim A, Saito S, Burnett J, et al. Population-Based HIV Impact Assessments Survey Methods, Response, and Quality in Zimbabwe, Malawi, and Zambia. J Acquir Immune Defic Syndr. 2021;87.

15. Patel HK, Duong YT, Birhanu S, Dobbs T, Lupoli K, Moore C, et al. A Comprehensive Approach to Assuring Quality of Laboratory Testing in HIV Surveys: Lessons Learned From the Population-Based HIV Impact Assessment Project. J Acquir Immune Defic Syndr. 2021;87.

16. Weld ED. Limits of Detection and Limits of Infection: Quantitative HIV Measurement in the Era of U = U. Journal of Applied Laboratory Medicine. 2021;6(1).

17. Saito S, Duong YT, Metz M, Lee K, Patel H, Sleeman K, et al. Returning HIV-1 viral load results to participant-selected health facilities in national Population-based HIV Impact Assessment (PHIA) household surveys in three sub-Saharan African Countries, 2015 to 2016: J Int AIDS Soc. 2017;20.

18. Farahani M, Radin E, Saito S, Sachathep KK, Hladik W, Voetsch AC, et al. Population Viral Load, Viremia, and Recent HIV-1 Infections: Findings from Population-Based HIV Impact Assessments (PHIAs) in Zimbabwe, Malawi, and Zambia. J Acquir Immune Defic Syndr (1988). 2021;87.

19. Wang Y, Xu G, Huang YW. Modeling the load of SARS-CoV-2 virus in human expelled particles during coughing and speaking. PLoS One. 2020;15(10 October).

20. He X, Lau EHY, Wu P, Deng X, Wang J, Hao X, et al. Temporal dynamics in viral shedding and transmissibility of COVID-19. Nat Med. 2020;26(5).

21. Gómez YM, Barranco-Chamorro I, Castillo JS, Gómez HW. An Extension of the Fréchet Distribution and Applications. Axioms [Internet]. 2024;13(4). Available from: https://www.mdpi.com/2075-1680/13/4/253

22. Ramos PL, Louzada F, Ramos E, Dey S. The Fréchet distribution: Estimation and application - An overview. Journal of Statistics and Management Systems. 2020;23(3).

23. Kneib T, Silbersdorff A, Säfken B. Rage Against the Mean – A Review of Distributional Regression Approaches. Econom Stat. 2023;26.

24. Han WM, Law MG, Egger M, Wools-Kaloustian K, Moore R, McGowan C, et al. Global estimates of viral suppression in children and adolescents and adults on antiretroviral therapy adjusted for missing viral load measurements: a multiregional, retrospective cohort study in 31 countries. Lancet HIV. 2021;8(12).

25. Glass T, Myer L, Lesosky M. The role of HIV viral load in mathematical models of HIV transmission and treatment: a review. BMJ Glob Health. 2020 Jan;5(1):e001800–001800. eCollection 2020.

26. Minchella PA, Chipungu G, Kim AA, Sarr A, Ali H, Mwenda R, et al. Specimen origin, type and testing laboratory are linked to longer turnaround times for HIV viral load testing in Malawi. PLoS One. 2017;12(2).

